# Physiologically Based Pharmacokinetic Modeling of Acoziborole Supplements Clinical Drug–Drug Interaction Data and Predicts Time-dependent Interaction Duration Following Single-Dose Oral Treatment for African Sleeping Sickness

**DOI:** 10.64898/2026.11.25.26368957

**Authors:** Jean-Yves Gillon, Estelle Weinling, Swati Jaiswal, Séverine Blesson, Laura M. Shireman, David Wesche, Lisa Almond

## Abstract

Acoziborole is a novel single-dose oral treatment for *gambiense* human African trypanosomiasis (g-HAT). Acoziborole’s long half-life and complex, time-dependent effects on drug-metabolizing enzymes necessitate drug–drug interaction (DDI) assessment. A physiologically based pharmacokinetic (PBPK) model was developed to characterize acoziborole DDI potential in African patients with g-HAT and support product labeling recommendations. Model development used available physicochemical, *in vitro* binding and interaction parameters, and clinical pharmacokinetic data from healthy participants and participants with g-HAT. Clinical DDI data for cytochrome P450 (CYP) 3A4 and CYP2D6 substrates were used to assess and refine the PBPK model. Predicted PK was validated using independent clinical datasets. The model was applied to predict clinically untested scenarios, including time-dependent DDI with CYP3A4 and CYP2D6 probe substrates, and DDIs with CYP1A2, CYP2B6, CYP2C8, CYP2C9, CYP2C19, and BCRP substrates. The model adequately described acoziborole PK in healthy participants and participants with g-HAT. Simulations predicted strong CYP3A4 induction to ∼4 weeks, then moderate to ∼9 weeks and CYP2D6 clinically relevant inhibition for up to 3 months post-acoziborole administration. Clinically relevant exposure reductions were predicted for concomitantly administered CYP3A4 substrates, particularly during the first month post-acoziborole. Prolonged and clinically relevant exposure increases in sensitive CYP2D6 substrates were predicted, although DDI risk was lower and shorter for CYP2D6 substrates also metabolized by CYP3A4. Simulated interactions with other enzymes were weak-to-moderate, with minimal transporter-mediated effects. The findings supported discussions with regulatory agencies to inform duration-based risk management strategies and prescribing information for acoziborole, and co-administration with common medications in the target population.

## 1 Introduction

Acoziborole (AN5568 or SCYX-7158), a benzoxaborole-6-carboxamide, received a positive EU-M4all scientific opinion in February 2026 as the first single-dose oral treatment for human African trypanosomiasis caused by *Trypanosoma brucei gambiense* (g-HAT), covering first-stage (hemo-lymphatic), second-stage (meningo-encephalitic), and severe second-stage disease (≥100 white blood cells/μL with or without trypanosomes in cerebrospinal fluid) in adolescents aged ≥12 years weighing ≥40 kg and adults.^1^ A parasite mRNA processing inhibitor,^2^ acoziborole is approved as a single oral dose of three 320 mg tablets. It represents a significant advance over previous standard-of-care treatments involving complex regimens, intravenous infusions, intramuscular injections and/or invasive diagnostics.^3^ Transmitted by the tsetse fly (*Glossina* spp.), untreated HAT is usually fatal. Although ∼55 million people across sub-Saharan Africa are at risk, extensive control efforts have kept reported cases below 1000 since 2018.^4^ The simple oral acoziborole regimen could accelerate progress toward the World Health Organization’s goal of HAT elimination by 2030.^5^

Following oral administration of a single 960 mg dose to fasted participants with g-HAT aged ≥15 years, acoziborole was slowly absorbed with a median peak blood concentration observed 48 h after dosing, plateauing until 96 h post-dose.^6^ Mean acoziborole peak blood concentration (C_max_) was 11.6 µg/mL (coefficient of variation [CV] 28.9%) and the mean area under the concentration–time curve (AUC) was 5216 µg·h/mL (CV 29.6%) (corresponding to 14.3 µg/mL and 6440 µg·h/mL in plasma, respectively).^6^ The mean terminal half-life (t_1/2_) of acoziborole in g-HAT patients was 296 h (12.3 days).^6^

The fraction absorbed (fa) was estimated to be at least 70%.^7^ Acoziborole is a substrate of the transporter P-glycoprotein, but intestinal efflux via this pathway is not expected to play an important role *in vivo*.^6^ Plasma protein binding was high with a fraction unbound (fu) of 0.0216, and a blood-to-plasma ratio of 0.81, indicating limited distribution into red blood cells.^8^ In a human mass-balance study, acoziborole had predominant but slow biliary-fecal elimination with limited metabolism.^7^ Radioactivity was mainly excreted in feces (74.2% of the dose by day 60 post-dose) and to a lesser extent in urine (10.9% of the dose by day 16).^7^ Renal elimination of unchanged acoziborole was <1% of the dose.^7^

*In vitro* data showed acoziborole induction of cytochrome P450 (CYP) 3A4 and time-dependent inhibition of CYP2D6, as well as inhibition of CYP2C8 and induction of CYP1A2, CYP2B6, CYP2C8, CYP2C9, CYP2C19, and UGT1A1 (data on file, Table 1). *In vitro* inhibition of several transporters by acoziborole was reported, including breast cancer resistance protein (BCRP), organic anion transporting polypeptide (OATP) 1B1, OAT3, organic cation transporter (OCT) 2, and multidrug and toxin extrusion (MATE) transporters (Table 1). Static drug–drug interaction (DDI) predictions using *in vitro* data and C_max_ observed in patients showed a potential *in vivo* DDI risk for all the *in vitro* CYP interactions. For transporters, only the BCRP interaction was significant, and other transporters were excluded as per ICH M12 guidance. Physiologically based pharmacokinetic (PBPK) modeling was used to assess the DDI liability dynamically over time considering induction and mechanism-based inactivation. The findings were used to design a clinical DDI study in healthy participants using sensitive index substrates for CYP3A4 (midazolam) and CYP2D6 (dextromethorphan).^9^ *In vivo*, acoziborole showed strong CYP3A4 induction, reducing midazolam AUC by 92% and C_max_ by 85% and shortening midazolam t_1/2_, in addition to strong CYP2D6 inhibition, increasing dextromethorphan AUC 35-fold and C_max_ 13-fold and prolonging dextromethorphan t_1/2_.^9^

**TABLE 1.** Acoziborole PBPK model final parameters.

| Parameter | Value | Source |
|---|---|---|
| *Physicochemical properties* |  |  |
| Molecular weight (g/mol) | 367.11 | Internal data |
| logP | 3.51 | Internal data |
| Compound type | Acid | Internal data |
| pKa | 9.61 | Internal data |
| B:P | 0.81 | Data on file (401247-20151013-BPR) |
| fu (healthy participants) | 0.0216 | Data on file (Exp 152-10) |
| fu (patients) | 0.03 | Clinical study (OXA-02-HAT) |
| Main binding protein | HSA | Assumed |
| *Human absorption, dissolution, metabolism and elimination* |  |  |
| fu<sub>gut</sub> | 1 | Assumed |
| P<sub>app</sub> (10<sup>-5</sup> cm/s) | 4.2 | MDCK, data on file (Exp 027-09) |
| P<sub>app</sub> (10<sup>-5</sup> cm/s) | 6.7 | Calibrator, data on file (Exp 027-09) |
| P<sub>eff,human</sub> (10<sup>-4</sup> cm/s) | 1.742 | Predicted from P<sub>app</sub> data |
| Q<sub>gut</sub> (L/h) | 9.272 | Predicted from P<sub>app</sub> data |
| ka (h<sup>-1</sup>) | 0.04 | Optimized, recover observed t<sub>max</sub> and C<sub>max</sub> in clinical study (DNDi-OXA-001) |
| fa | 0.7 | Clinical study (DNDi-OXA-03-HAT) |
| V<sub>SS</sub> (L/kg) | Predicted | Simcyp Method 2 |
| Kp scalar | 0.138 | Optimized to recover V<sub>SS</sub> estimate derived from clinical study (DNDi-OXA-03-HAT) |
| CL/F (L/h) | 0.12 | Clinical study (DNDi-OXA-001) |
| Biliary CL<sub>int</sub> (µL/min/million cells) | 0.326 | Retrograde mode; CL/F (0.12 L/h) obtained from clinical study (DNDi-OXA-001) |
| fu<sub>inc</sub> | 1 | Assumed |
| CL<sub>R</sub> (L/h) | 0.002 | Clinical study (DNDi-OXA-03-HAT) |
| *Drug interaction parameters* |  |  |
| CYP2C8 | *K*<sub>i</sub> 31.8 µM<br>fu<sub>mic</sub> 0.76 | Data on file (CYP2139 R2I)<br>Predicted |
| CYP2D6 | *K*<sub>i</sub> 27 µM<br>fu<sub>mic</sub> 0.61<br>*K*<sub>app</sub> 19.2 µM<br>*k*<sub>inact</sub> 89.2 h<sup>−1</sup> | Data on file (CYP2139 R2I)<br>Predicted<br>Data on file (CYP2139 R2I)<br>Refined to recover observed data for DDI with dextromethorphan in clinical study (DNDi-OXA-07-HAT) |
| CYP1A2 | Ind<sub>max</sub> 3.00<br>IndC<sub>50</sub> 10.40 µM | Data on file (CYP2139 R2 Induction) |
| CYP2B6 | Ind<sub>max</sub> 4.82<br>IndC<sub>50</sub> 2.85 µM | Data on file (CYP2139 R2 Induction) |
| CYP2C8 | Ind<sub>max</sub> 4.61<br>IndC<sub>50</sub> 1.72 µM | Data on file (CYP2139 R2 Induction) |
| CYP2C9 | Ind<sub>max</sub> 2.33<br>IndC<sub>50</sub> NC | Data on file (CYP2139 R2 Induction) |
| CYP2C19 | Ind<sub>max</sub> 2.58<br>IndC<sub>50</sub> NC | Data on file (CYP2139 R2 Induction) |
| CYP3A4 | Ind<sub>max</sub> 7.26<br>IndC<sub>50</sub> 0.72 µM | Optimized to recover observed values from DDI with midazolam in clinical study (OXA-07-HAT) |
| UGT1A1 | Ind<sub>max</sub> 10.6<br>Ind<sub>max</sub> 1.82 (calibrated)<br>IndC<sub>50</sub> 3.0 µM<br>fu<sub>hep</sub> 0.552 | Data on file (KLC-SA24-02–IHH0111).<br>Calibrated to rifampicin (*in vitro*, *in vivo*)<br>Data on file (KLC-SA24-02–IHH0111).<br>Calculated mean of values from two methods<sup>a</sup> |
| BCRP | *K*<sub>i</sub> 61.6 µM | Data on file (CYP2139 R2E) |
| OATP1B1 | *K*<sub>i</sub> 29.4 µM | Data on file (CYP2139 R2E) |
| OAT3 | *K*<sub>i</sub> 17.0 µM | Data on file (CYP2139 R2E) |
| OCT2 (renal) | *K<sub>i</sub>* 22.1 µM | Data on file (CYP2139 R2E) |
| MATEs (renal) | *K*<sub>i</sub> 43.1 µM | Data on file (CYP2139 R2E) |

<sup>a</sup> Austin, et al. *Drug Metab Dispos* 33, 419-25 (2005); Kilford, et al. *Drug Metab Dispos* 36, 1194-7 (2008).
B:P, blood/plasma partition ratio; BCRP, breast cancer resistance protein; CL/F, apparent clearance of drug after extravascular administration; CL<sub>int</sub>, intrinsic clearance of drug in eliminating organ; CL<sub>R</sub>, renal clearance of drug; C<sub>max</sub>, maximum plasma concentration; CYP, cytochrome P450; fa, fraction absorbed; fu, fraction unbound in plasma; fu<sub>gut</sub>, fraction unbound in gut enterocytes; fu<sub>hep</sub>, fraction unbound in hepatocytes; fu<sub>inc</sub>, fraction unbound in incubation; fu<sub>mic</sub>, fraction unbound in microsomes; HSA, human serum albumin; IndC<sub>50</sub>, calibrated inducer concentration producing half maximal induction; Ind<sub>max</sub>, calibrated maximal fold induction over vehicle control; ka, absorption rate constant; *K*<sub>app</sub>, apparent inactivation constant (concentration supporting half maximal rate of time-dependent inhibition); *K*<sub>i</sub>, reversible inhibition constant; *k*<sub>inact</sub>, maximal rate constant for enzyme inactivation; Kp scalar, tissue-to-plasma partition coefficient scalar; logP, logarithm of the octanol-water partition coefficient (neutral compound); MATEs, multidrug and toxin extrusion transporters; MDCK, Madin-Darby canine kidney cells; NC, not calculated; OATP, organic anion transporting polypeptide; OCT, organic cation transporter; P<sub>app</sub>, apparent permeability coefficient; P<sub>eff,human</sub>, effective human jejunum permeability; pKa, acid dissociation constant; Q<sub>gut</sub>, effective blood flow, the gut; t<sub>max</sub>, time of maximum plasma concentration; V<sub>SS</sub>, volume of distribution at steady state.

Comorbidities are common in HAT patients, most notably malaria.^10^ Given the long half-life of acoziborole, together with the complexity of enzyme induction and inhibition, multiple, lengthy clinical DDI studies would be needed to investigate these interactions. PBPK modeling is an established approach for predicting DDIs accepted by regulatory agencies in lieu of clinical studies.^11–22^

This work describes a PBPK model developed to support acoziborole DDI prescribing information. The aim was to assess the magnitude and duration of acoziborole’s effects on exposure (AUC and C_max_) for CYP3A4 and CYP2D6 substrates, including potential co-medications, in adult African patients with g-HAT. Additionally, the model was used to predict likely DDI outcomes of acoziborole as a precipitant with sensitive substrates of CYP1A2, CYP2B6, CYP2C8, CYP2C9, CYP2C19 and BCRP. In this context, PBPK modeling was employed to integrate the available clinical and *in vitro* data to enable a quantitative assessment of precipitant DDI risk and its temporal profile, supporting evidence-based dosing recommendations in adult patients with g-HAT.

## 2 Methods

### 2.1 Model Development

Simcyp (version 22; Certara, Sheffield, UK) was used to develop the acoziborole PBPK model. The modeling strategy is summarized in Figure 1. The model was parameterized using acoziborole physicochemical properties, mechanistic predictions based on *in vitro* data, and clinically derived inputs from a first-in-human single ascending dose study in healthy participants (DNDi-OXA-001), a Phase II study in participants with g-HAT (DNDi-OXA-02-HAT), and a mass-balance study in healthy participants (DNDi-OXA-03-HAT), with the model refined using clinical DDI data (DNDi-OXA-07-HAT) (Table S1).^7–9, 23^ First-order absorption and full PBPK distribution models were used. Complete model inputs, derivations, assumptions, and parameter-optimization procedures are provided in Table 1 and Methods S1. The PBPK model in healthy African participants showed good agreement with observed data (DNDi-OXA-001), with PK parameters within 1.13-fold of observed values (Figure S1). The healthy adult model was adapted for African participants with g-HAT, incorporating an increase in acoziborole fu from 0.02 to 0.03 consistent with clinical observations of reduced acoziborole binding to human serum albumin (Methods S1).^23^ The final model in adult African participants with g-HAT showed good agreement with observed data (DNDi-OXA-02-HAT), with PK parameters within 1.07-fold of observed values (Figure S2).

**FIGURE 1.**
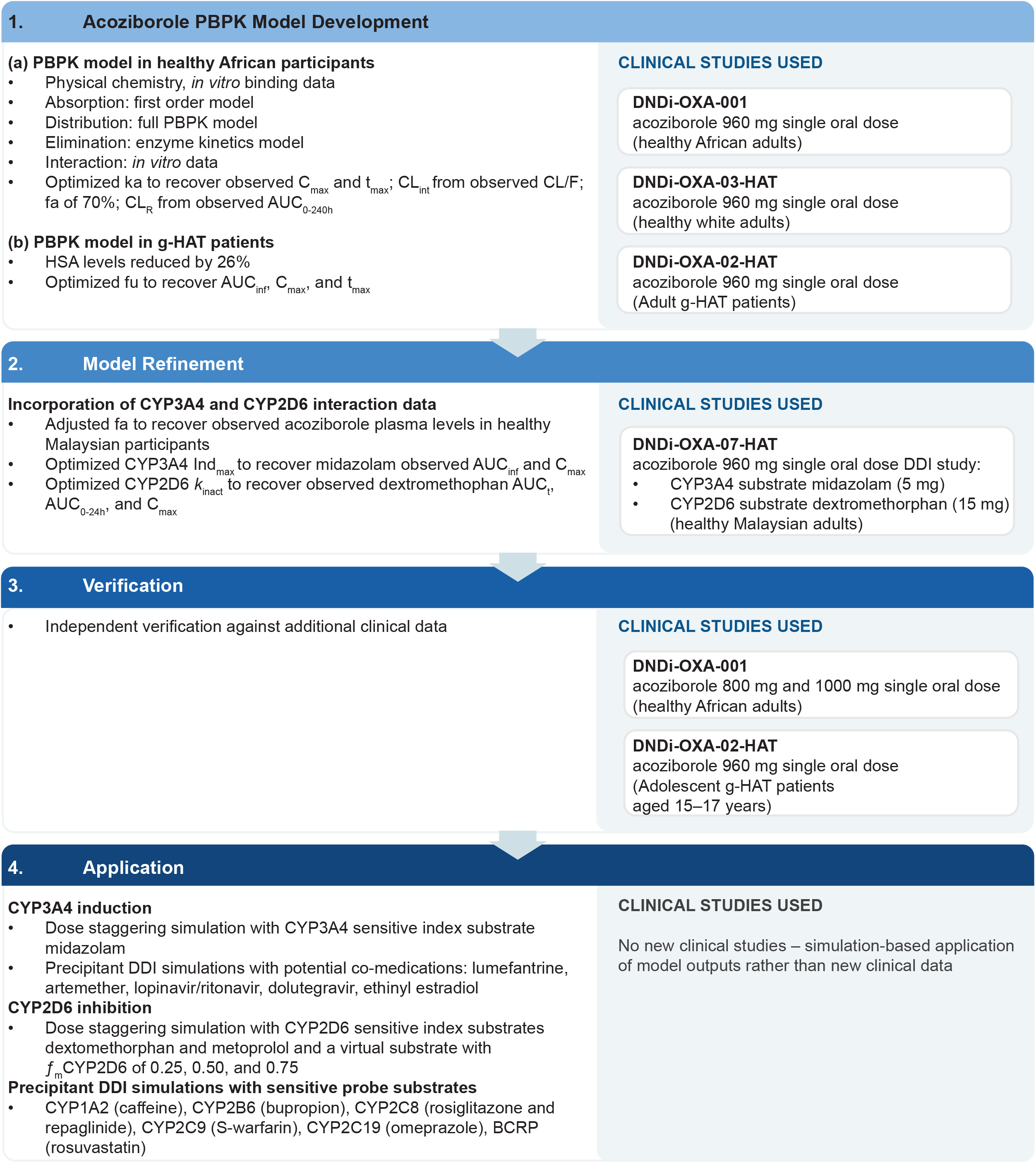
Modeling strategy. AUC, area under the concentration–time curve; AUC_inf_, AUC from time 0 extrapolated to infinity; AUC_t_, AUC from time 0 until last quantifiable concentration; AUC_0-24h_, AUC from time 0 to 24 h; AUC_0-240h_, AUC from time 0 to 240 h; BCRP, breast cancer resistance protein; CL/F, apparent clearance of drug after extravascular administration; CL_int_, intrinsic clearance of drug in eliminating organ; CL_R_, renal clearance of drug; C_max_, maximum plasma concentration; CYP, cytochrome P450; DDI, drug–drug interaction; fa, fraction absorbed; ƒ_m_, fraction metabolized; fu, fraction unbound in plasma; g-HAT, African trypanosomiasis caused by *Trypanosoma brucei gambiense*; HSA, human serum albumin; Ind_max_, calibrated maximal fold induction over vehicle control; ka, absorption rate constant; *k*_inact_, maximal rate constant for enzyme inactivation; t_max_, time of maximum plasma concentration.

### 2.2 Model Refinement

Model refinement to capture time-dependent enzyme induction and inhibition used clinical data from healthy Malaysian participants (study DNDi-OXA-07-HAT) (Figure 1).^9^ Simulations replicated the administration, with and without a single oral dose of acoziborole (960 mg), of the CYP3A4 sensitive index substrate midazolam (5 mg single dose) and CYP2D6 sensitive index substrate dextromethorphan (15 mg single dose). A sensitivity analysis approach was used to best recover observed clinical DDI results. *In vitro* CYP3A4 induction data overpredicted the magnitude of the DDI with midazolam, hence the maximal induction parameter (Ind_max_) was decreased from 12.1 to 7.26 (Methods S3). Although a strong DDI was predicted using the *in vitro* inactivation parameters, a 20-fold increase in the CYP2D6 maximal inactivation rate constant (*k*_inact_) from 4.46 h^-1^ to 89.2 h^-1^ was required to recover the observed DDI magnitude (Methods S3).

### 2.3 Simulated Populations

Simulations used a South African population library,^24^ aligned to clinical trial designs in healthy participants (DNDi-OXA-001) and African participants with g-HAT (DNDi-OXA-02-HAT).^23^ Height was underestimated and weight overestimated, so the input equations were modified to better recover the clinical population demographics (Methods S2). For adolescent DNDi-OXA-02-HAT participants, an African pediatric population^25^ was modified using published demographic data from pediatric patients with HAT.^26, 27^ Age–height and weight–height relationships were scaled by 0.95 and 0.85, respectively. For model refinement, simulations used the Simcyp healthy volunteer population matched to the clinical trial design (DNDi-OXA-07-HAT) in healthy male Malaysian participants.

### 2.4 Model Verification

Model performance was independently verified using additional clinical PK datasets for healthy African participants (DNDi-OXA-001) and participants with g-HAT (DNDi-OXA-02-HAT) (Figure 1).^8, 23^ Simulations were done for single oral doses of 800 mg and 1000 mg acoziborole in healthy African adults (male-only cohorts) using 10 virtual trials of 6 participants and for single oral doses of 960 mg acoziborole in adolescent patients with HAT (15–17 years; 55.6% female) using 10 trials of 20 participants. Model performance was assessed using predicted-to-observed ratios for AUC and C_max_.

### 2.5 Model Application

The refined acoziborole PBPK model was used to assess the strength and duration of DDIs with probe substrates and co-medications. Simcyp version 22 compound library models or previously verified PBPK models in the Simcyp model repository were used.^28, 29^ Strong, moderate, and weak interactions were defined as per ICH M12 guidance based on the ratios of geometric mean AUC and C_max_ in the presence of acoziborole divided by that in the absence of acoziborole (Table S2).^30^

#### 2.5.1 CYP3A4 Interaction

To estimate the duration of CYP3A4 induction, dose staggering DDI simulations were performed for the index substrate midazolam (5 mg single dose). The effect on exposure was assessed when the sensitive substrate was dosed on days 1, 2, 8, 15, 22, 29, 33, 36, 43, 50, 57, 64, 71, 85, 106 or 127 (acoziborole dosed on day 1).

Potential co-medications were selected from the WHO Essential Medicines List (Table S3). CYP3A4 substrate co-medication scenarios were simulated to predict exposure changes in clinical use settings. Simulations were done with or without acoziborole (single dose 960 mg) administered on day 1 for 10 trials of 10 African patients with g-HAT aged 18–68 years; 43.2% were female (except for ethinyl estradiol: 100% female). Simulations were done to include days 1 to 10 to reflect the point of maximum CYP3A4 induction following a single 960 mg acoziborole dose (Table S3).

The antimalarials lumefantrine and artemether were simulated using their standard and extended (recommended for co-administration with CYP3A4 inducers) regimens,^28, 31^ and with dosing delayed to day 36 post-acoziborole. For comparison to a standard strong CYP3A4 inducer, simulations were repeated for day 15 after multiple rifampicin doses (600 mg once daily). For lumefantrine, a clinically relevant concentration threshold of 280 ng/mL on day 7 post-dose was applied, as plasma concentrations below this level are associated with an increased risk of clinical failure.^32, 33^ DDI simulations were performed for ethinyl estradiol and the anti-HIV drugs dolutegravir and lopinavir/ritonavir using their standard dosing regimens. For UGT1A1 substrates (ethinyl estradiol, dolutegravir), *in vitro* induction parameters or values calibrated to rifampicin were considered (Table 1).

#### 2.5.2 CYP2D6 Interaction

To determine the duration of CYP2D6 inhibition, dose staggering DDI simulations were performed for dextromethorphan (15 mg single dose), metoprolol (100 mg single dose), and virtual compounds based on metoprolol, but with fraction metabolized (ƒ_m_) for CYP2D6 of 25%, 50%, and 75%. The effect on exposure was assessed when the sensitive substrate was dosed on days 1, 8, 29, 43, 64, 85, 92, 120, 127, 169 or 183 (acoziborole dosed on day 1).

#### 2.5.3 DDI Simulations with Sensitive Probe Substrates

The model was used prospectively to predict likely DDI outcomes of acoziborole as a precipitant for induction of index substrates of CYP1A2 (caffeine), CYP2B6 (bupropion), CYP2C9 (*S*-warfarin), and CYP2C19 (omeprazole), induction or inhibition of CYP2C8 index substrates (rosiglitazone and repaglinide), and inhibition of BCRP (rosuvastatin) (Table S4). Object dosing was timed to maximize the magnitude of the interaction (Table S4). To account for uncertainty in acoziborole non-specific binding and evaluate the ‘worst-case’ scenario for DDI, simulations were repeated for CYP inhibition with a 10-fold reduced enzyme competitive inhibition constant for unbound drug (*K*_i,u_),^34^ CYP induction with a 10-fold increase in Ind_max_, and BCRP inhibition with *K*_i,u_ values reduced by 15-fold.^35^ Sensitivity analyses were performed adjusting parameters for the minor pathway to examine effects caused solely by that enzyme or transporter.

## 3 Results

### 3.1 Acoziborole Physiologically Based Pharmacokinetic Model

Using the final model (Table 1), simulated single-dose acoziborole profiles at 800 mg and 1000 mg in healthy African participants were consistent with clinical data (Figure S3), with geometric mean AUC_inf_ and C_max_, and median t_max_ within 1.6-fold of observed values (Table S5). For African patients with g-HAT, simulations in adolescents (≥15 to <18 years) showed good agreement with observed clinical data (Figure S3), with PK parameters within 1.34-fold of observed values (Table S5).

### 3.2 CYP3A4 Interaction

The duration of the CYP3A4 interaction following a single 960 mg acoziborole dose is summarized in Figure 2. Post-acoziborole, the decrease of midazolam (ƒ_m_CYP3A4 97.8%) exposure was maximal when dosed on day 8 and strong when dosed up to day 33, moderate between days 36 and 64, weak from day 71, and negligible by day 126 (Table S6).

**FIGURE 2.**
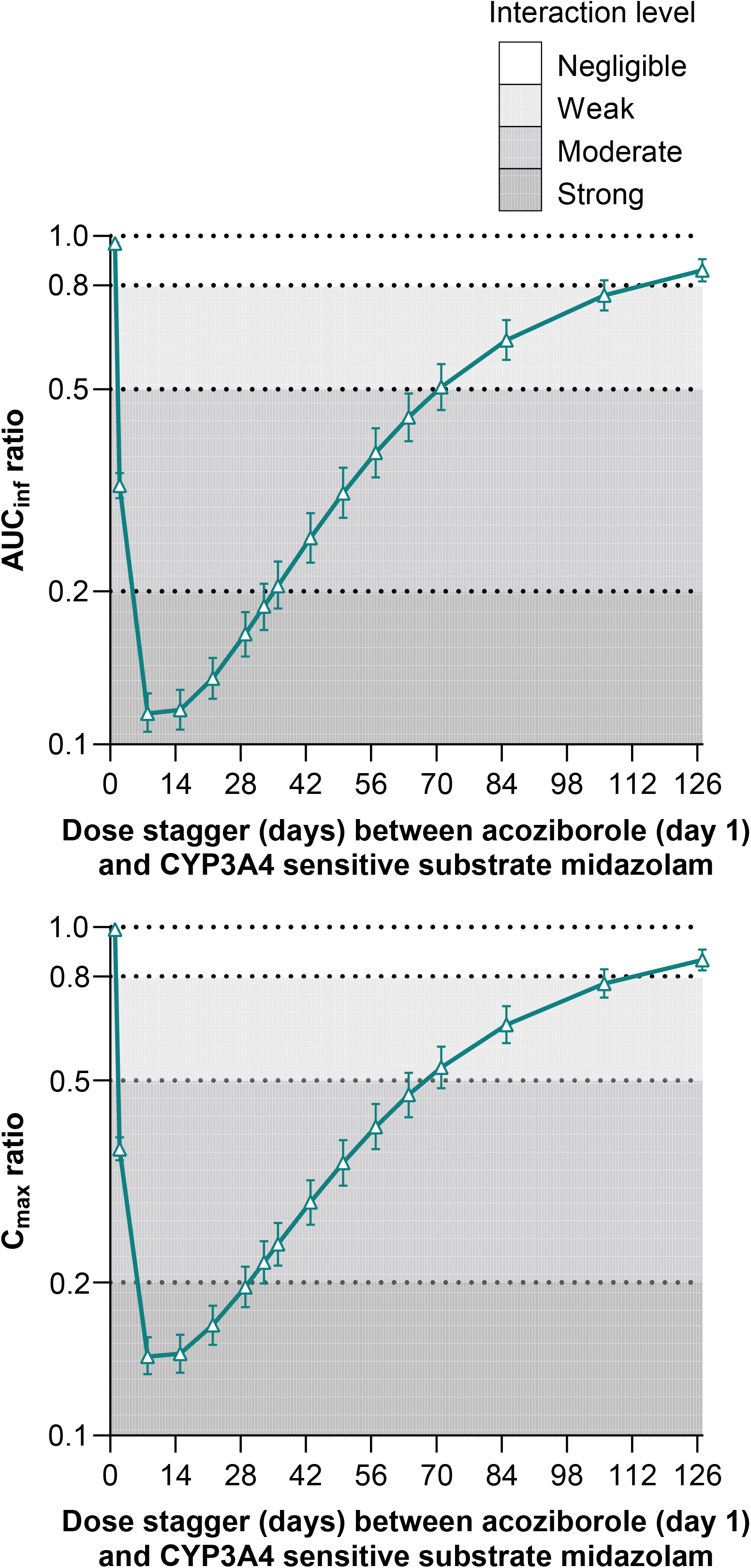
CYP3A4 duration of interaction. Simulated plasma geometric mean AUC_inf_ and C_max_ ratios and geometric 90% CIs (with acoziborole : without acoziborole) for a single dose of midazolam (5 mg) when co-administered at varying times after a single dose of 960 mg acoziborole in African adult participants with g-HAT. AUC_inf_, area under the plasma concentration–time curve from time 0 extrapolated to infinity; C_max_, maximum plasma concentration; CYP, cytochrome P450; Ind_max_, calibrated maximal fold induction over vehicle control.

Simulations were done to predict DDIs with potential co-medications. For the standard 3-day regimen of the antimalarial lumefantrine, mean simulated plasma concentrations were reduced with acoziborole administration. Post-acoziborole, there was a strong interaction when lumefantrine was dosed at day 10 and a moderate interaction when dosed at day 36 (Figure 3A, Table S7). The proposed therapeutic threshold (>280 ng/mL at 168 h)^32, 33^ was not met in simulations of the standard 3-day lumefantrine regimen dosed on either day 10 or day 36 post-acoziborole or for the extended 5-day regimen^31^ when dosed on day 10 post-acoziborole. However, simulated lumefantrine concentrations reached the threshold for clinical efficacy with the extended 5-day regimen when initiated at day 36 post-acoziborole (Figure S4). The magnitude of the interaction and partial exposure recovery with the extended regimen were similar for acoziborole and for rifampicin (Figure S4, Table S7).

**FIGURE 3.**
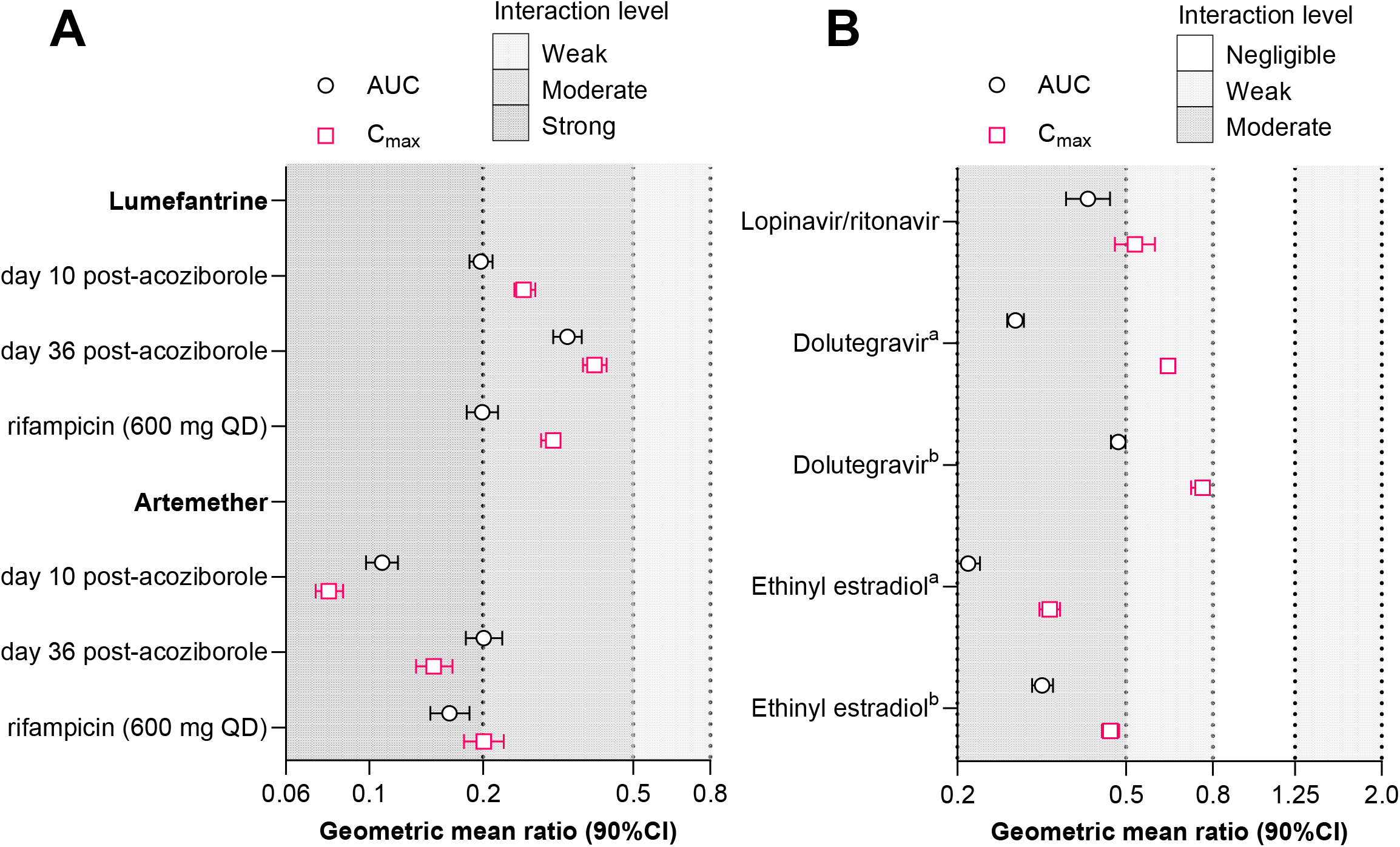
Forest plot of simulated plasma geometric mean AUC and C_max_ ratios for potential co-medications. A) Lumefantrine and artemether at their standard regimens starting on day 10 or day 36 following a single dose of 960 mg acoziborole or in the presence of multiple doses of rifampicin (600 mg once daily [QD]). For day 10 dosing AUC_216-672h_, and for day 36 AUC_840-1296h_ with or without acoziborole and AUC_336-792h_ with or without rifampicin. B) Potential co-medications administered at their standard regimens at the time of maximum CYP3A4 induction or CYP2D6 inhibition following a single dose of 960 mg acoziborole. ^a^ Simulations using *in vitro* UGT1A1 Ind_max_; ^b^ Simulations using UGT1A1 Ind_max_ calibrated to rifampicin (Table 1). Lopinavir AUC_216-240h_, dolutegravir AUC_216-240h_, ethinyl estradiol AUC_216-744h_. AUC, area under the concentration–time curve; C_max_, maximum plasma concentration; CYP, cytochrome P450.

The antimalarial artemether isa CYP3A4 and CYP2B6 substrate,and a strong decrease in exposure waspredicted with the standard regimen administered on days 10 and 36 post-acoziborole (Figure 3A, Table S8). The extended artemether regimen did not fully recover the equivalent exposures seen with the standard regimen in the absence of acoziborole (Figure S5). A similar extent of interaction was simulated with multiple rifampicin doses (Table S8).

Two anti-HIV drugs were assessed (Figure 3B, Table S9). Clinically, lopinavir is co-administered with ritonavir to enhance exposure via CYP3A4 inhibition. Co-administration with acoziborole diminished this effect via CYP3A4 induction, resulting in a net moderate interaction and decreasing lopinavir exposure (Figure S6). For dolutegravir, there was also a moderate effect on exposures (Figure S7), which tended to be greater when the *in vitro* UGT1A1 Ind_max_ was used versus when calibrated to rifampicin (Figure 3B, Table S9).

For the oral contraceptive ethinyl estradiol, there was a moderate reduction in exposure with acoziborole, which tended to be greater when using uncalibrated *in vitro* UGT1A1 Ind_max_ values rather than calibrating to rifampicin (Figure 3B, Table S9, Figure S8).

### 3.3 CYP2D6 Interaction

The duration of the acoziborole CYP2D6 interaction following a single 960 mg acoziborole dose is summarized in Figure 4. When dosing with dextromethorphan (ƒ_m_CYP2D6 97.3%), a strong increase in exposure was predicted, with a maximal effect on day 29 and a moderate effect from day 85 until day 127, post-acoziborole (Table S10). A moderate increase in exposure was predicted for metoprolol (ƒ_m_CYP2D6 66.8%; ƒ_m_CYP3A4 17.3%) administered until day 92 and a weak increase from day 120 to 127 post-acoziborole (Table S11). As anticipated for the virtual compounds, increasing the ƒ_m_CYP2D6 from 25% to 75% increased the strength and duration of the interaction (Table S12).

**FIGURE 4.**
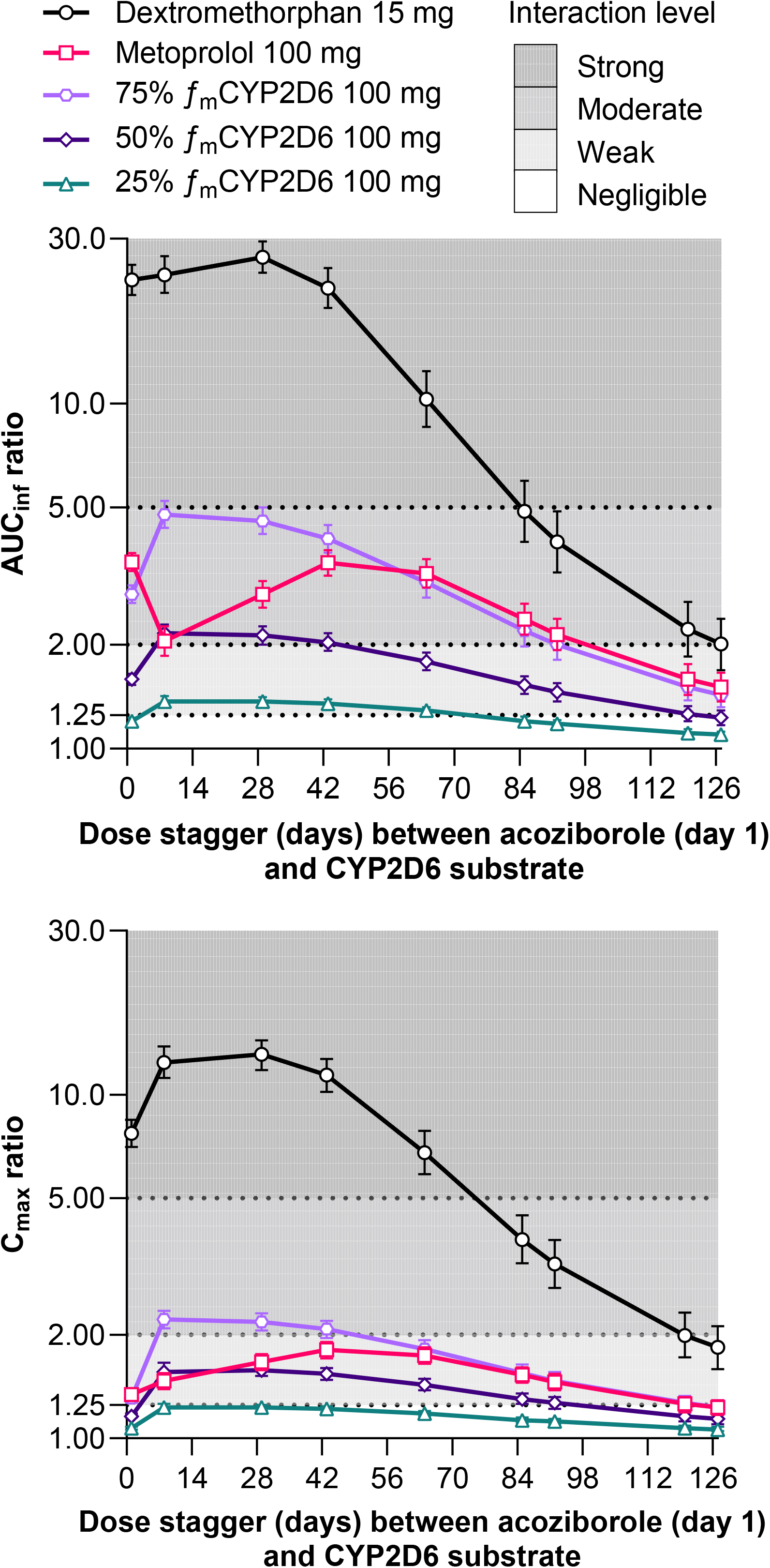
CYP2D6 duration of interaction. Simulated plasma geometric mean AUC_inf_ and C_max_ ratios and geometric 90% CIs (with acoziborole : without acoziborole) for sensitive substrates when co-administered at varying times after a single dose of 960 mg acoziborole in adult African participants with g-HAT. CYP2D6 substrates: a single dose of dextromethorphan (15 mg), or a single dose of metoprolol (100 mg) and hypothetical substrates with a fraction metabolized (ƒ_m_) for CYP2D6 of 25%, 50%, and 75%. AUC_inf_, area under the plasma concentration–time curve from time 0 extrapolated to infinity; C_max_, maximum plasma concentration; CYP, cytochrome P450; Ind_max_, calibrated maximal fold induction over vehicle control.

### 3.4 DDI Simulations with Sensitive Probe Substrates

Using *in vitro*-derived interaction parameters and scheduling administration of each sensitive substrate at the timepoint predicted to yield the maximal DDI, PBPK modeling indicated predominantly weak-to-moderate effects of a single 960 mg acoziborole dose on exposure (Figure 5, Table S13). Weak effects on exposures were predicted for rosuvastatin (BCRP substrate; 1.25-fold increase in AUC), caffeine (CYP1A2 substrate; 27% decrease in AUC) and bupropion (CYP2B6 substrate; 43% decrease in AUC). Moderate exposure reductions were predicted for *S*-warfarin (CYP2C9 substrate; 51% decrease in AUC), rosiglitazone (CYP2C8/CYP2C9 substrate; 55% decrease in AUC), repaglinide (CYP2C8/CYP3A4 substrate; 65% decrease in AUC) and omeprazole (CYP2C19 substrate; 67% decrease in AUC). These findings are consistent with existing evidence that enzymes regulated by the pregnane X receptor (CYP2C8, CYP2C9, and CYP2C19) exhibit lower inducibility relative to CYP3A4.^30^

**FIGURE 5.**
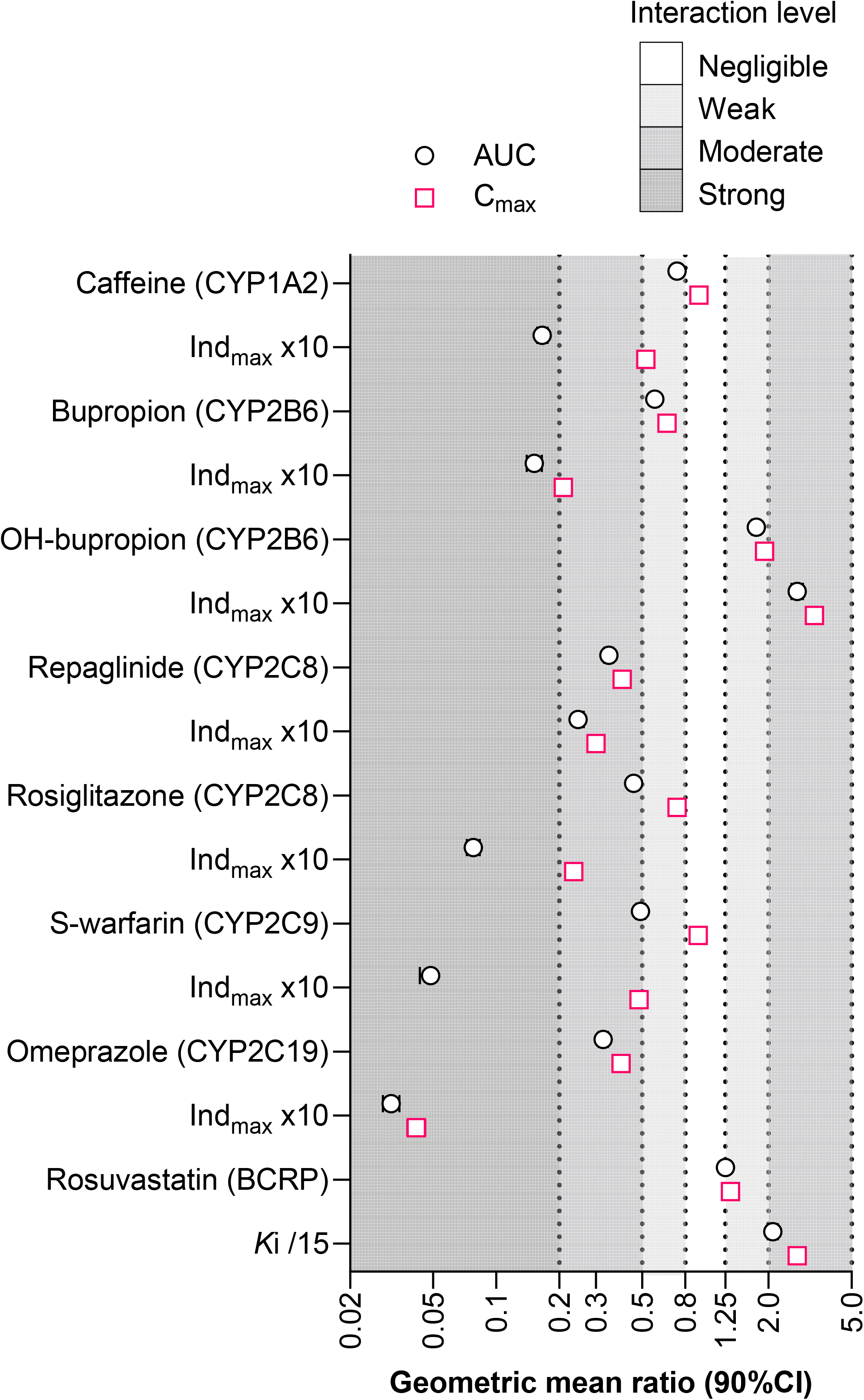
Forest plot comparing plasma AUC and C_max_ ratios for objects of DDI probe substrates when co-administered with a single dose of 960 mg acoziborole. Predicted AUC and C_max_ are shown for the cytochrome P450 enzyme substrates using the *in vitro* Ind_max_ and for a 10-fold increase in Ind_max_ as a sensitivity analysis. For the BCRP transporter rosuvastatin, *in vitro K*_i_ was used and a 15-fold reduction in *K*_i_ as a sensitivity analysis. Additional sensitivity analyses are shown in Table S14. AUC, area under the concentration–time curve – for repaglinide, rosiglitazone, and omeprazole AUC extrapolated to infinity (AUC_inf_) was used and for caffeine, bupropion, OH-bupropion, rosuvastatin, and S-warfarin, AUC until the time of last quantifiable concentration (AUC_t_) was used; BCRP, breast cancer resistance protein; C_max_, maximum plasma concentration; CYP, cytochrome P450; Ind_max_, calibrated maximal fold induction over vehicle control; *K*_i_, reversible inhibition constant.

Sensitivity analyses indicated that the predicted magnitude of DDIs increased to moderate for the BCRP substrate rosuvastatin and remained moderate for the CYP2C8/CYP3A4 substrate repaglinide. For rosiglitazone, both CYP2C8 inhibition and induction effects were included in the model and the sensitivity analysis indicated induction to be the primary interaction (Table S14). Increasing Ind_max_ 10-fold increased the interaction with acoziborole to strong for substrates of CYP1A2 (caffeine), CYP2B6 (bupropion), CYP2C8 (rosiglitazone), CYP2C9 (S-warfarin), and CYP2C19 (omeprazole) (Figure 5, Table S14).

## 4 Discussion

This work integrated *in vitro* data, clinical PK data in healthy participants and participants with g-HAT (the target population), a clinical DDI study in healthy participants, and PBPK modeling to define the time course and magnitude of acoziborole as a perpetrator of DDIs. The results support a mechanistic understanding of acoziborole as a strong CYP3A4 inducer, a strong time-dependent CYP2D6 inhibitor, and a moderate-to-weak inducer of multiple additional CYP enzymes (CYP2C8, CYP2C9, CYP2C19, CYP1A2, CYP2B6) and UGT1A1, with minimal clinically relevant transporter inhibition. Importantly, the PBPK framework enabled translation of these effects into a time-varying interaction risk profile (Figure 6).

**FIGURE 6.**
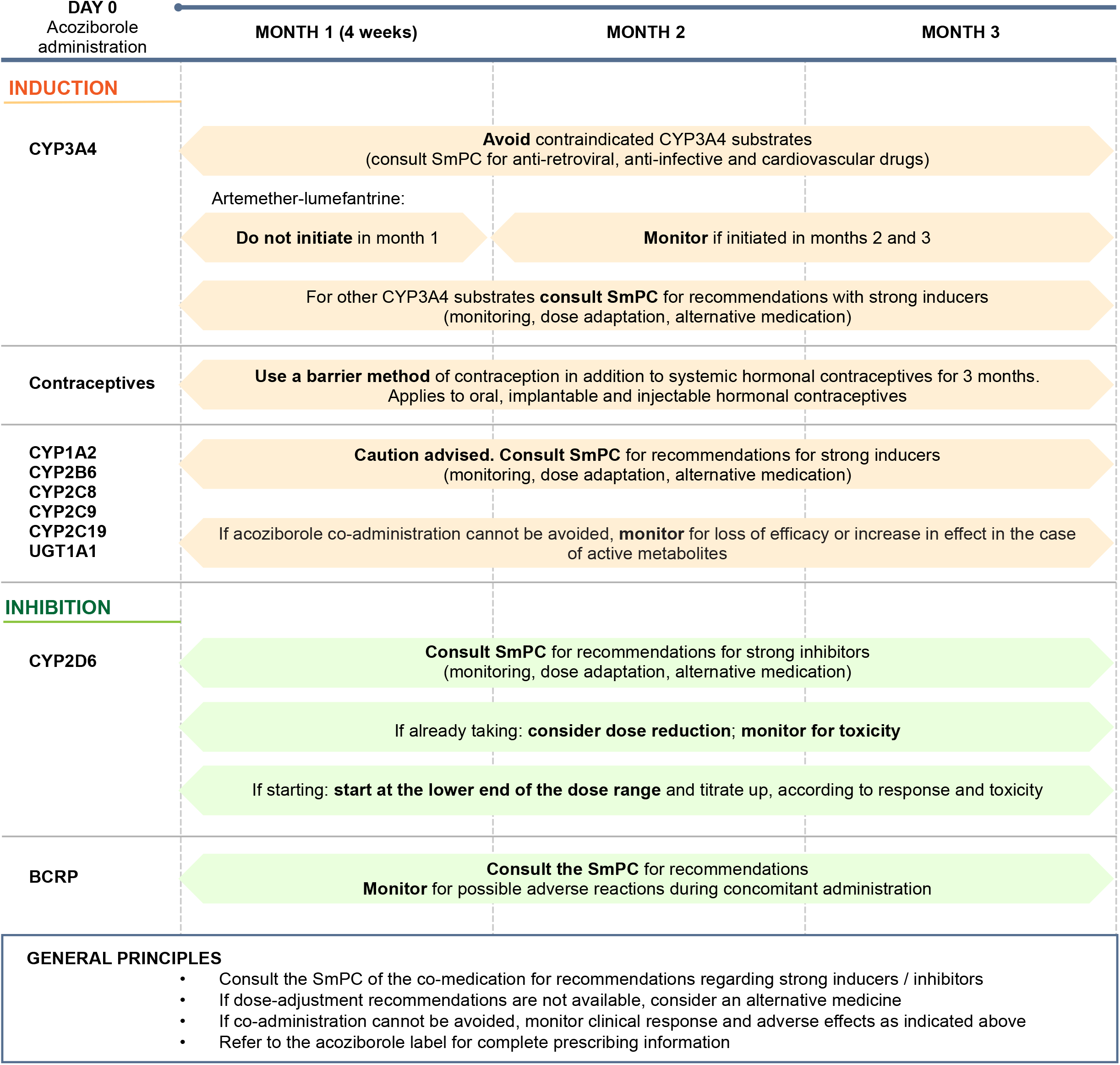
Summary of anticipated acoziborole drug–drug interaction risk translated into time-based recommendations for co-medications relevant to g-HAT patients. BCRP, breast cancer resistance protein; CYP, cytochrome P450; SmPC, summary of product characteristics (prescribing information).

A key outcome of the PBPK analysis was the delineation of the time-dependent nature of DDI risk following a single therapeutic acoziborole oral dose (960 mg), driven by prolonged elimination and the turnover kinetics of affected enzymes. CYP3A4 induction was predicted to peak approximately 1 week post-dose and to remain strong for up to 4 weeks, moderate for up to 9 weeks with residual effects extending to around 3 months. CYP2D6 inhibition lasted for approximately 3 months post-acoziborole dose. The results for dextromethorphan, with a ƒ_m_CYP2D6 close to 100%, provide information on the maximal effect expected on a sensitive CYP2D6 substrate with a high ƒ_m_CYP2D6 (>0.95), representing a worst-case scenario with a high and sustained interaction. Acoziborole may produce a moderate increase up to approximately 3 months post-dose with CYP2D6 substrates with ƒ_m_CYP2D6 >0.75 and with CYP2D6 substrates also metabolized by CYP3A4, such as metoprolol.^36^ For other substrates with an ƒ_m_CYP2D6 of around 0.75 or lower, the DDI magnitude was less (weak to moderate). Many CYP2D6 substrates, such as metoprolol, are also CYP3A4 substrates. For these substrates, CYP3A4 induction offsets the CYP2D6 inhibition to some degree and hence the predicted DDI related to CYP2D6 is reduced. Thus, the overall DDI risk is governed by overlapping enzyme modulation profiles, supporting duration-based risk management recommendations that encompass the period of maximal and residual enzyme effects following acoziborole administration. In the prescribing information, based on these predictions, caution was advised for 3 months after acoziborole administration for CYP2D6 and CYP3A4 substrates, to cover the duration of exposure to acoziborole and the time required to recover CYP activities after acoziborole elimination.

In the clinical studies in participants with g-HAT, concomitant medications were permitted without DDI-specific restrictions. Exposure to CYP enzyme substrates affected by acoziborole was limited, and no associated safety signal or clinically apparent loss of efficacy was identified.^1, 23^ Malaria is common in g-HAT–endemic regions and frequently requires prompt treatment with antimalarials that are CYP3A4 substrates.^37^ All reported malaria episodes treated with standard-dose artemether–lumefantrine resolved clinically in patients treated on days −1 to 7 (*n*=16), days 8–30 (*n*=5), and day ≥31 (*n*=46) relative to acoziborole administration.^1, 23^ In accordance with local guidelines, cure was assessed clinically, with parasitological testing reserved for suspected treatment failure. These observations are, therefore, supportive but do not exclude reduced antimalarial exposure or subclinical treatment failure. Additional evidence was provided by the PBPK simulations, which predicted the greatest reduction in artemether– lumefantrine exposure during the first month after acoziborole administration. Accordingly, artemether–lumefantrine is contraindicated during the first month following acoziborole administration, but is permitted with caution thereafter, when CYP3A4 induction is predicted to be reduced (Figure 6). Clinical experience and conservative PBPK predictions together supported the risk-based labeling approach, without additional clinical DDI studies.

Simulations for oral ethinyl estradiol showed reduced exposure following acoziborole. In line with recommendations for other strong CYP3A4 inducers, women using hormonal contraceptives should use an additional reliable barrier method or use an alternative contraception method for up to 3 months after acoziborole administration (Figure 6).

Simulations were done comparing the effects of acoziborole with rifampicin, showing broadly similar magnitudes of CYP3A4 induction. These findings supported rifampicin as a clinically relevant benchmark for risk communication and labeling of acoziborole CYP3A4-mediated interactions. The acoziborole prescribing information, therefore, considers drugs that are contraindicated with rifampicin or strong CYP3A4 inducers.^6^ Accordingly, co-administration recommendations for antiretroviral therapies were aligned with existing rifampicin guidance. Ritonavir-boosted protease inhibitors, including atazanavir, darunavir, and lopinavir are contraindicated for up to 3 months following acoziborole administration, reflecting the risk of sustained reduction in exposure.^6^ In contrast, integrase inhibitors such as dolutegravir and raltegravir (primarily metabolized by UGT1A1), and efavirenz (primarily metabolized by CYP2B6), may be used with caution or dose-adaptation, particularly during the first 4 weeks after acoziborole treatment, as recommended in their prescribing information with rifampicin, considering potential variability in net exposure.^6, 38^

This rifampicin framework can be extended to other CYP3A4 substrates commonly used in endemic settings. For example, praziquantel, an antiparasitic agent frequently used in regions co-endemic for helminth infections, is contraindicated for up to 3 months after acoziborole dosing, consistent with existing contraindications for use with strong and moderate CYP3A4 inducers.^39^ Overall, leveraging established rifampicin-based guidance provides a pragmatic and clinically interpretable approach to managing acoziborole DDIs, while accounting for its prolonged and time-dependent CYP3A4 induction profile.

The acoziborole PBPK model indicates a distinct interaction profile characterized by concurrent CYP3A4 induction and strong CYP2D6 inhibition, in contrast to prototypical inducers such as rifampicin that act predominantly through induction alone. This dual mechanism has important implications for predicting DDIs, as the net effect on exposure depends on the relative contribution of each metabolic pathway and their time-dependent interplay. For example, substrates highly dependent on CYP2D6, such as dextromethorphan, were predicted to experience more pronounced and prolonged increases in exposure, whereas drugs metabolized via multiple pathways, including both CYP2D6 and CYP3A4, such as metoprolol, showed more moderate overall effects due to CYP inhibition and induction. This interplay becomes particularly relevant for many concomitant medications used in g-HAT–endemic settings, including antipsychotics, antidepressants, beta-blockers and analgesics, which are often cleared through multiple enzyme pathways. In such cases, the magnitude and direction of interaction may be attenuated or less predictable, although simulations suggest that even for compounds with CYP2D6 contributions of up to approximately 75%, the overall clinical impact was generally modest. The acoziborole PBPK model, therefore, provides a valuable framework for the systematic evaluation of these clinically relevant multi-pathway interactions, enabling more informed risk assessment and supporting context-appropriate guidance for co-administration in endemic settings.

Several limitations should be considered when interpreting the PBPK model. Although dose non-linearity was observed at lower doses, model development was based on clinical data at the therapeutic 960 mg oral single dose. Given the limited mechanistic understanding of this non-linearity and the lack of clinical relevance of lower exposures, model performance outside the 800–1000 mg range was not evaluated. Differences in pharmacokinetics between healthy African participants and participants with g-HAT were attributed to changes in the unbound fraction, consistent with reported alterations in human serum albumin in this population.

However, in the absence of direct measurements in patients, this assumption remains unverified. The use of a virtual South African population represents an additional source of uncertainty, although adjustments were made to better reflect the body composition of clinical populations from the Democratic Republic of Congo where clinical trials of participants with g-HAT were performed. Clinical data on DDI predictions are limited to acoziborole as a perpetrator for interactions with CYP3A4 and CYP2D6 substrates. For other metabolic pathways, the absence of *in vivo* data precludes model verification. While sensitivity analyses were conducted to explore worst-case scenarios, the overall uncertainty remains, particularly for predicting complex interactions involving concurrent inhibition and induction or the contribution of transporters. However, the clinical DDI study anchored predictions of CYP3A4 as the enzyme most sensitive to induction. Hence, co-administration recommendations are conservative and did not require further clinical assessment to support labeling.

PBPK approaches have been used to predict CYP- and transporter-mediated interactions for both new and approved compounds to evaluate clinically untested precipitant and object of DDI scenarios to support regulatory recommendations.^11–22^ Acoziborole presents a distinctive challenge because of its exceptionally long elimination half-life and time-dependent effects on enzyme activity. The current study demonstrates how a mechanistic framework can complement clinical DDI data by characterizing the time course and persistence of enzyme induction and inhibition during sustained drug exposure. The PBPK model robustly characterized acoziborole as a strong CYP3A4 inducer and time-dependent CYP2D6 inhibitor, and allowed exploratory investigation of CYP2C8, CYP2C9, CYP2C19, CYP1A2, CYP2B6, UGT1A1, and transporter interactions. By quantifying the temporal evolution of CYP3A4 and CYP2D6 interactions and translating these predictions into duration-based co-medication recommendations, this work informed regulatory discussions and provided clinically actionable information where DDI studies would have been challenging.

## Study Highlights

- What is the current knowledge on the topic?

- A clinical drug–drug interaction (DDI) study evaluating a single 960 mg oral dose of acoziborole showed strong CYP3A4 induction and strong CYP2D6 inhibition, supported by *in vitro* enzyme and transporter interaction data.
- What question did this study address?

- Given acoziborole’s long half-life, the extent and duration of potentially complex, time-dependent DDIs across multiple metabolic pathways were evaluated to inform evidence-based recommendations for co-medication dose adjustments.
- What does this study add to our knowledge?

- A validated PBPK model enabled predictions of the extent and duration of CYP3A4 induction and CYP2D6 inhibition and the evaluation of exposure of sensitive index substrates of BCRP, CYP1A2, CYP2B6, CYP2C8, CYP2C9, and CYP2C19 in the presence of acoziborole.
- How might this change clinical pharmacology or translational science?

- Combined with clinical and *in vitro* data, PBPK model DDI predictions supported discussions with regulators, informing acoziborole prescribing information and clinical co-administration guidance. This scalable approach illustrates how PBPK modeling can complement clinical DDI studies and efficiently address data gaps for long-acting agents where DDIs reflect systemic exposure and prolonged enzyme modulation.

## Acknowledgements

Naomi Richardson (Magenta Communications Ltd, Abingdon, UK) wrote the first draft of this article based on internal reports and provided editorial and graphic assistance, funded by DNDi. The authors thank Nada Abla (MMV Medicines for Malaria Venture) for helpful discussion and acknowledge the support of Ping Zhao (Gates Foundation).

## Author contributions

Designed Research: J-YG, EW, SJ, LA, SB; Performed Research: SJ, LMS, LA; Analyzed Data: All authors; Contributed New Reagents/Analytical Tools: SJ, LA; Wrote manuscript: J-YG, EW, LA; All authors revised the manuscript.

## Data availability

Interested researchers may request access to de-identified participant data from Vivli, the data-sharing partner of the Drugs for Neglected Diseases initiative (DNDi), commissioner of this study, at https://vivli.org/ourmember/dndi/.

## Ethics

All clinical studies contributing data to this analysis were approved by the relevant independent ethics committees or institutional review boards and conducted in accordance with ICH Good Clinical Practice, applicable regulatory requirements, and the Declaration of Helsinki. Written informed consent was obtained from adult participants and from the parents or legal guardians of minors, with participant assent obtained where appropriate.

## Supplementary Information

Additional methods and results.

