## Supplementary Information for "Physiologically Based Pharmacokinetic Modeling of Acoziborole Supplements Clinical Drug–Drug Interaction Data and Predicts Time-dependent Interaction Duration Following Single-Dose Oral Treatment for African Sleeping Sickness"

**Table S1: Acoziborole clinical study data used for PBPK model development, refinement, and verification.**

| Clinical study | ClinicalTrials.gov registration | Description | Application to model |
|---|---|---|---|
| DNDi-OXA-001 | NCT01533961 | Determination of the optimal single dose treatment for acoziborole, a novel drug for the treatment of human African trypanosomiasis: first-in-human study | **Development:** adult healthy African males, acoziborole 960 mg single oral dose<br>**Verification:** adult healthy African males, acoziborole 800 and 1000 mg single oral doses |
| DNDi-OXA-02-HAT | NCT03087955 | Efficacy and safety of acoziborole in patients with human African trypanosomiasis caused by *Trypanosoma brucei gambiense*: a multicenter, open-label, single-arm, phase 2/3 trial | **Development:** adult African participants with g-HAT, acoziborole 960 mg single oral dose<br>**Verification:** adolescent (15 to 17 years) participants, acoziborole 960 mg single oral dose |
| DNDi-OXA-03-HAT | NCT04270981 | Mass balance, pharmacokinetics, metabolism, and excretion of radiolabeled acoziborole, a potential novel treatment for human African trypanosomiasis, following single microtracer oral dose, humans | **Development:** ADME data for model parameterization; adult healthy White males, acoziborole 960 mg single oral dose |
| DNDi-OXA-07-HAT | NCT05947604 | DDI study of single oral dose of acoziborole with sequential co-administration of midazolam and dextromethorphan | **Refinement:** healthy adult Malaysian males, acoziborole 960 mg single oral dose |

ADME, absorption, distribution, metabolism, excretion; DDI, drug–drug interaction; g-HAT, African trypanosomiasis caused by *Trypanosoma brucei gambiense*.

**Methods S1: Acoziborole PBPK model parametrization**

Distribution models were tested with a full PBPK model selected. Steady-state volume of distribution ($V_{SS}$) predicted using Rodgers and Rowland (*Pharm Res*. 2007;24:918-933)[1] was inconsistent with observed preclinical and population PK estimates. Instead, $V_{SS}$ was inferred, assuming complete intestinal and hepatic availability and a known fraction absorbed (fa 0.7). This yielded a $V_{SS}$ of 0.478 L/kg, and tissue partition coefficients (Kp) were subsequently scaled to recover this value.

With negligible *in vitro* metabolic turnover, intrinsic clearance ($CL_{int}$) was inferred via retrograde calculation from observed oral clearance (CL/F 0.12 L/h) (DNDi-OXA-001), yielding a total unbound intrinsic hepatic clearance ($CLu_{int,H}$) of 3.90 L/h, which was converted to a hepatocyte intrinsic clearance using liver weight and hepatocellularity factors, yielding a $CL_{int}$ of 0.326 µL/min/million cells. Low metabolic turnover and high permeability resulted in the fraction escaping first-pass metabolism approaching unity. The fraction unbound in enterocytes ($fu_{gut}$) was set to 1; a conservative assumption for the purpose of predicting DDIs, that all intracellular drug was available for metabolism. *In vitro* parameters describing enzyme inhibition and induction (notably CYP2D6 and CYP3A4) and transporter interactions were integrated.

***Optimization of absorption rate constant (ka).***

The effect of ka on acoziborole pharmacokinetic parameters was investigated in ten virtual trials of 6 healthy African participants (0% female) aged 26 to 44 years (as in clinical study DNDi-OXA-001). Simulated acoziborole $AUC_{inf}$, CL/F, $C_{max}$, and $t_{max}$ values were compared to observed data from Clinical Study DNDi-OXA-001 after a single 960 mg oral dose of acoziborole in healthy participants in the fasted state (table below). The ka value of 0.04 $h^{-1}$ best recovered the observed $C_{max}$ and $t_{max}$ and best described the observed plasma concentration–time profile on visual inspection (figure below).

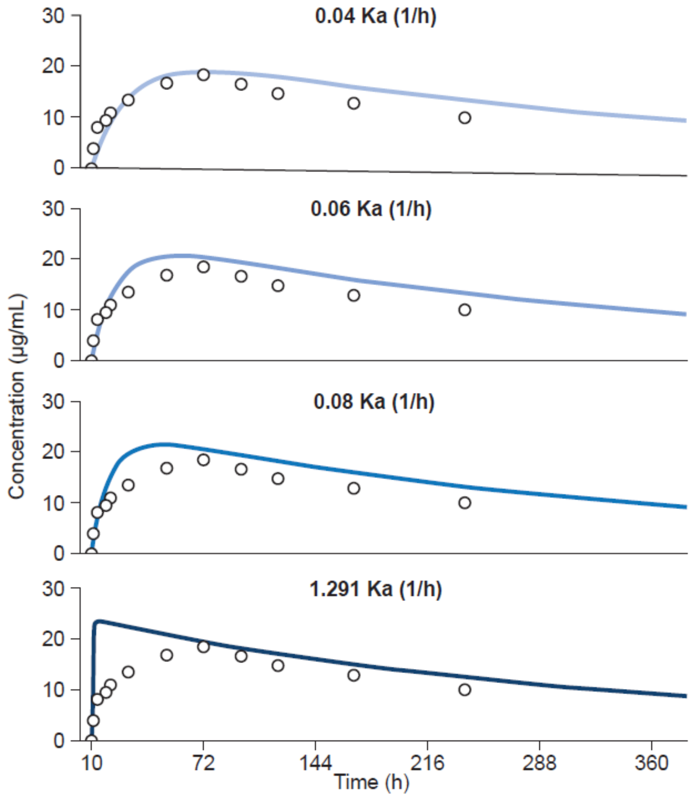

[1] Rodgers T, Rowland M. Mechanistic approaches to volume of distribution predictions: understanding the processes. Pharm Res. 2007;24:918–933.

| ka values | Population | $AUC_{inf}$ (µg/mL·h) | CL/F (L/h) | $C_{max}$ (µg/mL) | $t_{max}$ (h) |
|---|---|---|---|---|---|
| 1.29 $h^{-1}$ | Simulated | 9015 | 0.107 | 22.9 | 3.50 |
| | Observed | 7979 | 0.120 | 19.2 | 72.0 |
| | Simulated/observed | 1.13 | 0.889 | 1.19 | 0.049 |
| 0.08 $h^{-1}$ | Simulated | 9015 | 0.107 | 20.4 | 45.28 |
| | Observed | 7979 | 0.120 | 19.2 | 72.0 |
| | Simulated/observed | 1.13 | 0.889 | 1.06 | 0.629 |
| 0.06 $h^{-1}$ | Simulated | 9015 | 0.11 | 19.8 | 56.1 |
| | Observed | 7979 | 0.120 | 19.2 | 72.0 |
| | Simulated/observed | 1.13 | 0.889 | 1.03 | 0.778 |
| 0.04 $h^{-1}$ | Simulated | 9015 | 0.106 | 18.2 | 75.8 |
| | Observed | 7979 | 0.120 | 19.2 | 72.0 |
| | Simulated/observed | 1.13 | 0.885 | 0.949 | 1.05 |

Values are geometric mean, except $t_{max}$ which is the median. $AUC_{inf}$, area under the plasma concentration–time curve extrapolated to infinity; CL/F, apparent plasma clearance after extravascular administration; $C_{max}$, maximum plasma concentration; ka, absorption rate constant; $t_{max}$, time to maximum plasma concentration.

***Acoziborole PBPK model in African g-HAT patients; impact of infection on fraction unbound.***

The model was adapted for participants with g-HAT using the Simcyp South African population library adjusted to align simulated body size to the clinical population (18–68 years; 43.2% female) (study DNDi-OXA-02-HAT). Simulations were based on adult HAT participant data with 10 virtual trials of 20 participants.

As acoziborole is highly bound to plasma proteins, changes in human serum albumin (HSA), as reported in the literature and observed in clinical data from previous studies in HAT patients with fexinidazole studies (an oral HAT therapy), were considered a potential mechanism underlying the observed pharmacokinetic differences. Based on these data, a 26% reduction in HSA was incorporated into the model to represent African participants with HAT, resulting in an increase in the unbound fraction (fu) from 0.02 to 0.03 in the simulated population. The simulated HSA levels were consistent with those observed clinically in participants with HAT (study DNDi-OXA-02-HAT). However, this adjustment alone was insufficient to recover the magnitude of the clinically observed changes in pharmacokinetics. Previous experience with PBPK modeling in other disease settings has shown that alterations in plasma protein levels do not always fully account for changes in unbound fraction (fu). Therefore, the fu input value in the model was further optimized from 0.0216 to 0.03. This, alongside the changes in HSA, yielded a fu value of 0.04 in the simulated population and enabled the model to adequately capture the observed clinical pharmacokinetics in HAT patients (table below).

| Values for fu input into the model | Simulated population fu | Population | $AUC_{inf}$ (µg/mL·h) | CL/F (L/h) | $C_{max}$ (µg/mL) | $t_{max}$ (h) |
|---|---|---|---|---|---|---|
| 0.0216 | 0.03 | Simulated | 6430 | 0.149 | 15.6 | 71.0 |
| | | Observed | 4975 | 0.193 | 11.0 | NA |
| | | Simulated/observed | 1.29 | 0.774 | 1.42 | |
| 0.03 | 0.04 | Simulated | 4669 | 0.206 | 11.7 | 69.9 |
| | | Observed | 4975 | 0.193 | 11.0 | NA |
| | | Simulated/observed | 0.938 | 1.07 | 1.06 | |

Values are geometric mean, except $t_{max}$ which is the median. $AUC_{inf}$, area under the blood concentration-time curve extrapolated to infinity; CL/F, apparent blood clearance of drug after extravascular administration; $C_{max}$, maximum blood concentration; $t_{max}$, time to maximum blood concentration.

**Methods S2: Simulated populations.**

Demographic data were available for acoziborole studies in healthy adults of African origin (DNDi-OXA-001); adult and adolescent participants with g-HAT living in the Democratic Republic of Congo (DRC) (DNDi-OXA-02-HAT) and healthy participants living in Malaysia (DNDi-OXA-07-HAT). Additionally, demographic data from African pediatric participants with g-HAT receiving fexinidazole or Nifurtimox-Eflornithine Combination Therapy (NECT) were used to develop a representative adolescent population (table below).

| Study type | Study number | N (% female) | Age, years | Participants | Simulated population | Simulations |
|---|---|---|---|---|---|---|
| Single ascending dose study | DNDi-OXA-001 | 102 (0%) | 18–<45 | Healthy African participants | Modified South African healthy population[1] | Healthy African adult participants aged 18–44 years; 0% female |
| Phase II (≥18 years of age) | DNDi-OXA-02-HAT | 199 (43.2%) | ≥18–<69 | African participants with g-HAT | Modified South African healthy population[1] | Adult g-HAT patients aged 18–68 years; 43.2% female |
| Phase II (<18 years of age) | DNDi-OXA-02-HAT | 9 (55.6%) | ≥15–<18 | African participants with g-HAT | Modified African pediatric population[2] | Adolescent g-HAT patients aged 15–17; 55.6% female |
| NECT | NECT field study | 151 (41.1%) | 1–18 | African participants with g-HAT | | |
| Fexinidazole phase II | DNDiHATFEX006 | 125 (46.4%) | 6–14 | African participants with g-HAT | | |
| DDI study | DNDi-OXA-07-HAT | 19 (0%) | 20–35 | Healthy Malaysian participants | Modified healthy volunteer population | Healthy Malaysian adult participants aged 20–35 years; 0% female |

DDI, drug–drug interaction; g-HAT, African trypanosomiasis caused by *Trypanosoma brucei gambiense*.

For matching simulated and observed subject demographics, simulations of 20 trials of 20 participants each of the South African population were performed. Height tended to be underestimated, and weight tended to be overestimated, and the input equations were modified to better recover the clinical population demographics. This modified population was used for simulations in African participants with g-HAT.

The African pediatric population was used to simulate demographics of African pediatric participants with g-HAT for two independent clinical studies (NECT field study and DNDiHATFEX006) with age, height and weight matched to the respective study designs. Weight tended to be overestimated for both studies. By changing the age-to-height relationship by a factor of 0.95 and the weight-to-height equation by a factor of 0.85, the body sizes of African participants with g-HAT were better represented. This population library was then used to simulate the body sizes of adolescent participants in clinical study DNDi-OXA-02-HAT, with reasonable agreement to the observed data.

The Simcyp healthy volunteer library was used to simulate demographics for Malaysian healthy participants in clinical study DNDi-OXA-07-HAT using a matched trial design. Weight and height were overestimated compared to the clinical data, and the input equations were modified to better recover the clinical population demographics.

[1] Humphries, H. *et al.* Development of physiologically-based pharmacokinetic models for standard of care and newer tuberculosis drugs. *CPT Pharmacometrics Syst Pharmacol* **10**, 1382-95 (2021).

[2] Pan, X. & Rowland Yeo, K. Physiologically Based Pharmacokinetic Modeling to Determine the Impact of CYP2B6 Genotype on Efavirenz Exposure in Children, Mothers and Breastfeeding Infants. *Clin Pharmacol Ther* **114**, 182-91 (2023).

**Figure S1: Acoziborole PBPK model in healthy African participants.**

The acoziborole model in healthy African participants showed good agreement with the observed data (DNDi-OXA-001). Simulation is for acoziborole 960 mg single dose. Grey lines represent the 5th and 95th percentiles and the black line the arithmetic mean data for the simulated population (n = 60; 10 trials of 6 participants per trial). Individual observed data are shown as circles. Top is linear plot and bottom panel is log–linear plot.

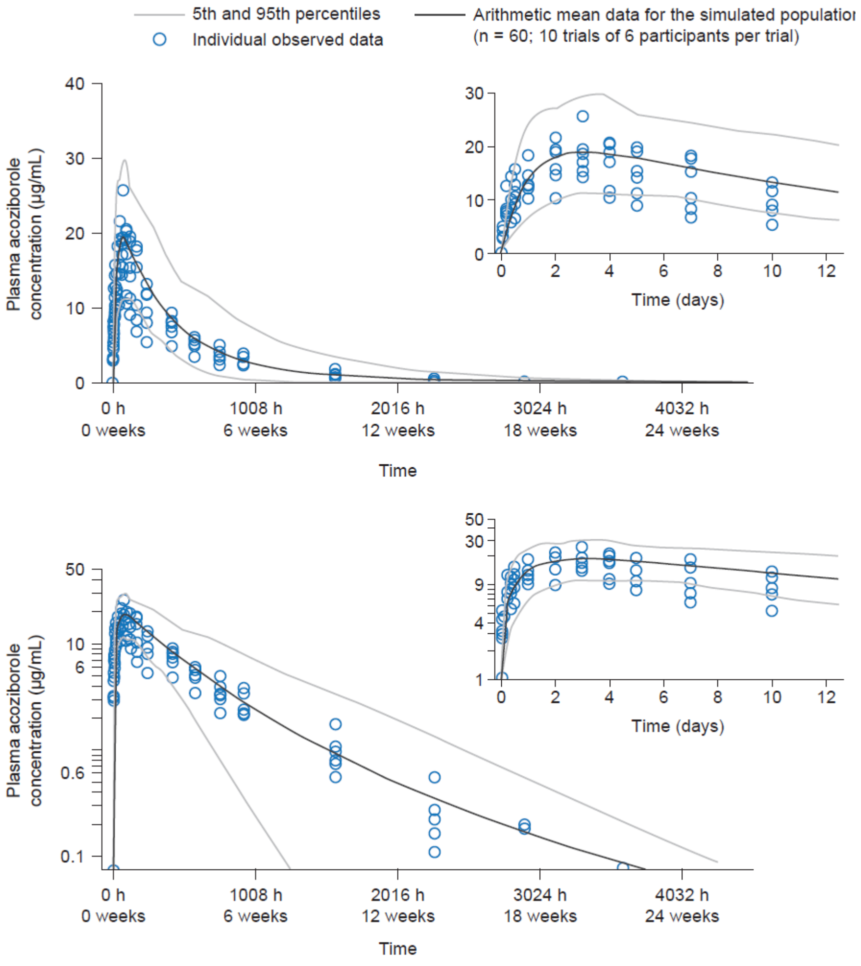

| Statistic | AUC$_{0\text{-inf}}$ (µg/mL·h) | CL/F (L/h) | C$_{max}$ (µg/mL) | t$_{max}$ (h) |
|---|---|---|---|---|
| Simulated | 9015 | 0.106 | 18.2 | 75.8 |
| CV% | 47.0 | 47.0 | 32.8 | – |
| 90% CI | 8186, 9927 | 0.0967, 0.117 | 17.0, 19.5 | 44.2, 126 |
| Observed | 7979 | 0.120 | 19.2 | 72.0 |
| Simulated/observed | 1.13 | 0.885 | 0.949 | 1.05 |

Values are geometric mean, except $t_{max}$ which is the median. AUC$_{0\text{-inf}}$, area under the plasma concentration–time curve from time 0 extrapolated to infinity; CI, confidence interval; CL/F, apparent clearance of drug after extravascular administration; $C_{max}$, maximum plasma concentration; CV%, coefficient of variation; $t_{max}$, time of maximum plasma concentration.

**Figure S2: Acoziborole PBPK model in African participants with g-HAT.**

The final model in African participants with g-HAT showed good agreement with the observed data (DNDi-OXA-02-HAT). Simulation is for acoziborole 960 mg single dose. Grey lines represent the 5th and 95th percentiles and the black line the arithmetic mean data for the simulated population (n = 200; 10 trials of 20 participants per trial). Individual observed data are shown as circles. Top is linear plot and bottom panel is log–linear plot.

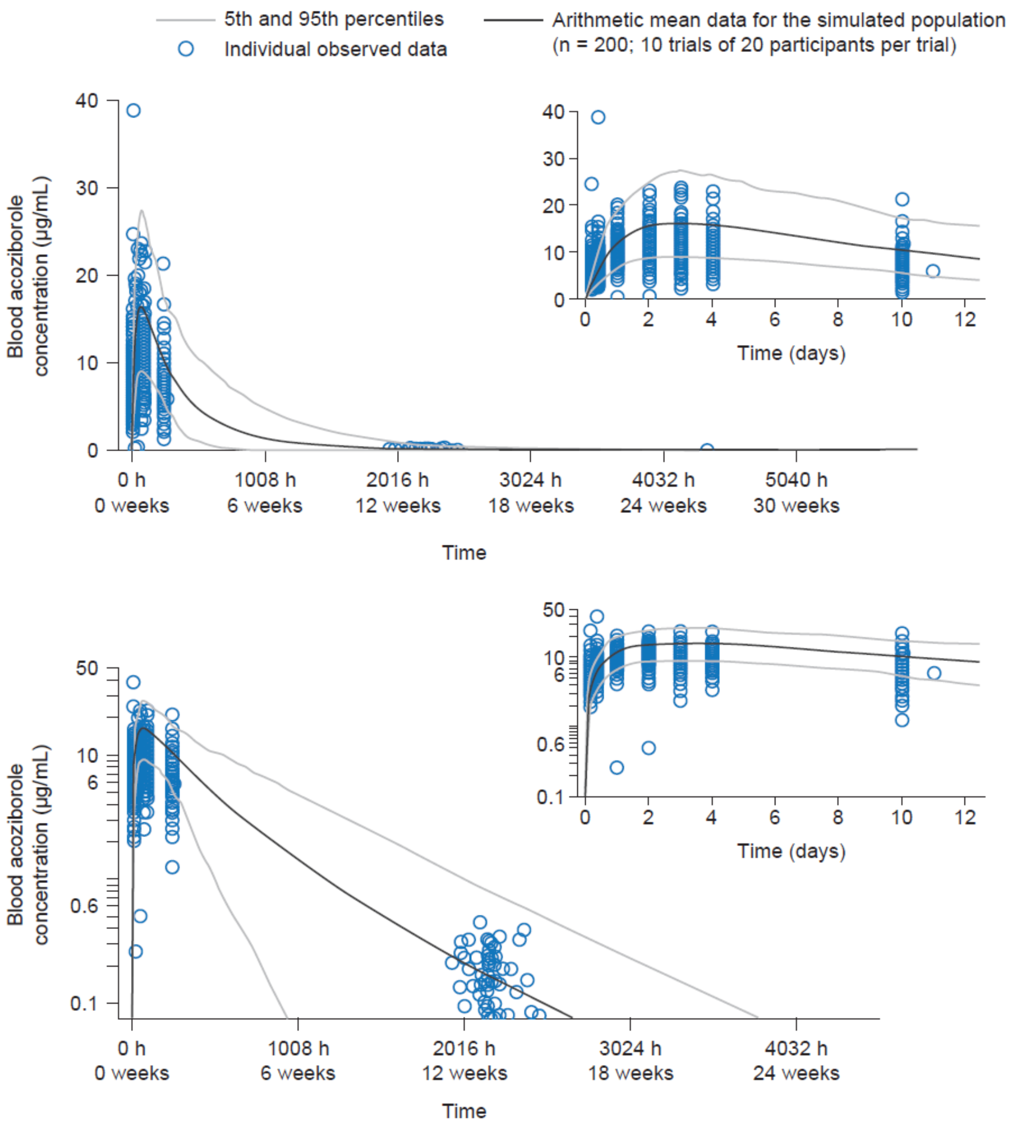

| Statistic | $AUC_{0\text{-inf}}$ (µg/mL·h) | CL/F (L/h) | $C_{max}$ (µg/mL) | $t_{max}$ (h) |
|---|---|---|---|---|
| Simulated | 4669 | 0.206 | 11.7 | 69.9 |
| CV% | 49.9 | 49.9 | 35.7 | – |
| 90% CI | 4419, 4933 | 0.195, 0.217 | 11.2, 12.1 | 39.8, 120 |
| Observed | 4975 | 0.193 | 11.0 | NA |
| Simulated/observed | 0.938 | 1.07 | 1.06 | – |

Values are geometric mean, except $t_{max}$ which is the median. $AUC_{0\text{-inf}}$, area under the blood concentration–time curve from time 0 extrapolated to infinity; CI, confidence interval; CL/F, apparent clearance of drug after extravascular administration; $C_{max}$, maximum blood concentration; CV%, coefficient of variation; $t_{max}$, time of maximum blood concentration.

**Methods S3: Model refinement for CYP3A4 induction and CYP2D6 inhibition using a clinical DDI study in healthy Malaysian participants (DNDi-OXA-07-HAT).**

***Recovery of observed acoziborole PK in healthy Malaysian participants.***

To recover the observed PK (DNDi-OXA-07-HAT), modification of the healthy participant acoziborole PBPK model was required with the fraction absorbed (fa) adjusted from 0.7 to 0.6 (figure below).

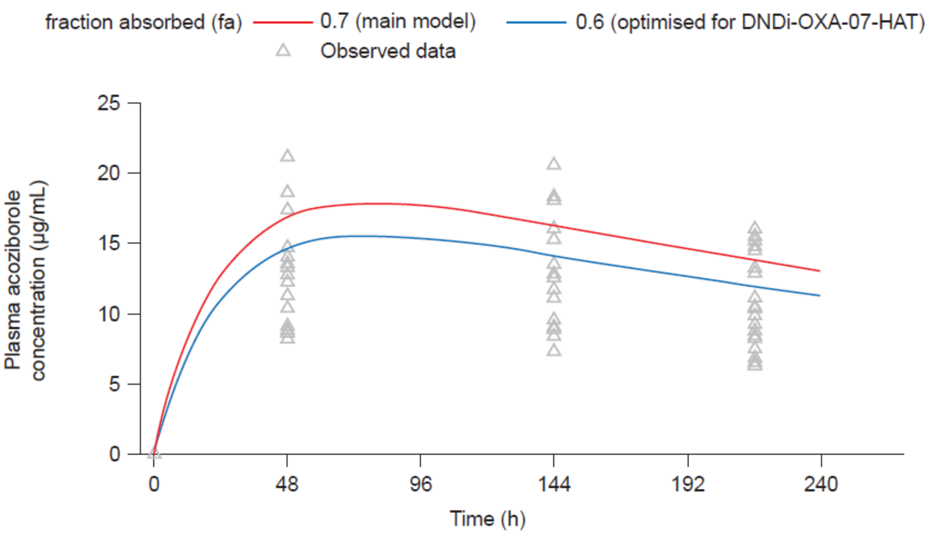

***CYP3A4 induction: Simulation of single oral dose of 5 mg midazolam co-administered with a single oral dose of 960 mg acoziborole in healthy participants.***

Using the above PBPK model, the *in vitro* CYP3A4 induction parameters ($Ind_{max}$ 12.10; $IndC_{50}$ 0.72 µM) underpredicted the observed midazolam geometric mean $AUC_{inf}$ and $C_{max}$ from the clinical study DNDi-OXA-07-HAT. $Ind_{max}$ was refined via an automated sensitivity analysis with $Ind_{max}$ of 12.10 and 10-fold lower and $IndC_{50}$ of 0.72 µM and 10-fold higher (figure below).

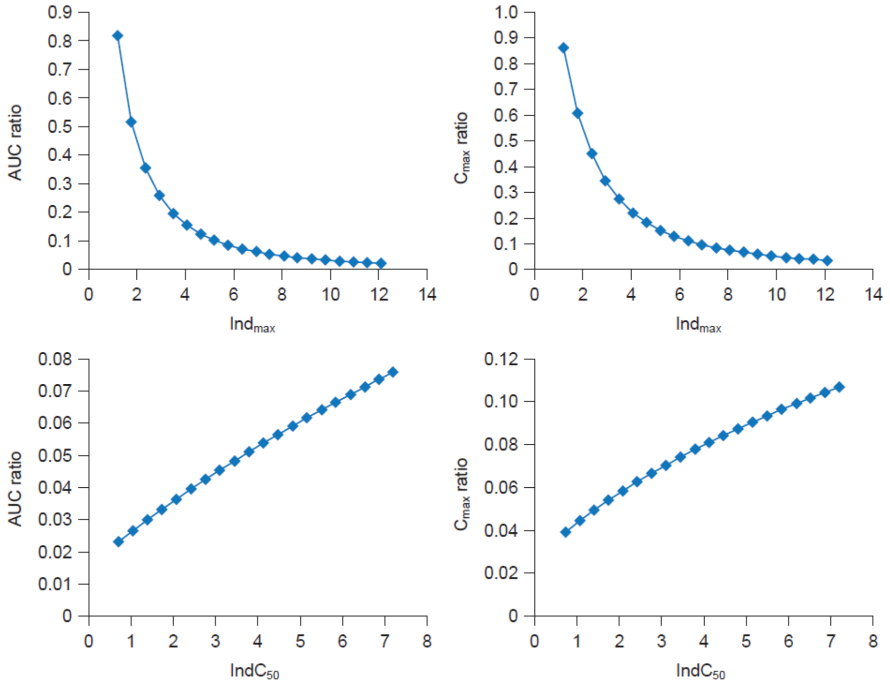

$IndC_{50}$, calibrated inducer concentration producing half maximal induction; $Ind_{max}$, calibrated maximal fold induction over vehicle control.

$Ind_{max}$ was a more sensitive parameter than $IndC_{50}$ and a manual sensitivity analysis was done decreasing $Ind_{max}$ by 1.66-fold and increasing $IndC_{50}$ by 7-fold versus *in vitro* estimates. Ten virtual trials of 19 healthy participants (0% female) aged 20 to 35 years as in clinical study (DNDi-OXA-07-HAT) were generated for a 5 mg midazolam dose administered on day 10 after a single dose of 960 mg acoziborole. An $Ind_{max}$ value of 7.26 provided the best agreement with the observed midazolam exposure, returning simulated geometric mean ratios within 1.15-fold of observed values (table below). Accordingly, an $Ind_{max}$ of 7.26 was applied in subsequent simulations and $IndC_{50}$ was unchanged from 0.72 µM.

| Model | Statistic | Without acoziborole | | With acoziborole | | Geometric mean ratio | |
|---|---|---|---|---|---|---|---|
| | | $AUC_{inf}$ (ng/mL·h) | $C_{max}$ (ng/mL) | $AUC_{inf}$ (ng/mL·h) | $C_{max}$ (ng/mL) | $AUC_{inf}$ ratio | $C_{max}$ ratio |
| CYP3A4 $Ind_{max}$ = 12.1 (*in vitro* value) | Simulated | 56.0 | 20.2 | 2.27 | 1.28 | 0.0405 | 0.0633 |
| | CV% | 86.2 | 73.5 | 110 | 112 | 69.2 | 66.0 |
| | 90% CI | 51.2, 61.3 | 18.7, 21.9 | 2.04, 2.52 | 1.15, 1.42 | 0.0375, 0.0436 | 0.0589, 0.068 |
| | Observed | 102.5 | 28.8 | 8.73 | 4.43 | 0.085 | 0.154 |
| | Simulated/observed | 0.546 | 0.702 | 0.260 | 0.289 | 0.475 | 0.411 |
| CYP3A4 $Ind_{max}$ = 7.26 (optimized) | Simulated | 56.0 | 20.2 | 5.33 | 2.81 | 0.0951 | 0.139 |
| | CV% | 86.2 | 73.5 | 105 | 102 | 59.6 | 54.9 |
| | 90% CI | 51.2, 61.3 | 18.7, 21.9 | 4.80, 5.91 | 2.53, 3.11 | 0.0890, 0.102 | 0.131, 0.148 |
| | Observed | 102.5 | 28.8 | 8.73 | 4.43 | 0.085 | 0.154 |
| | Simulated/observed | 0.546 | 0.702 | 0.610 | 0.633 | 1.12 | 0.903 |
| CYP3A4 $Ind_{C50}$ = 5.04 µM | Simulated | 56.0 | 20.2 | 5.22 | 2.56 | 0.093 | 0.127 |
| | CV% | 86.2 | 73.5 | 111 | 104 | 62.7 | 55.9 |
| | 90% CI | 51.2, 61.3 | 18.7, 21.9 | 4.68, 5.81 | 2.31, 2.83 | 0.087, 0.100 | 0.119, 0.135 |
| | Observed | 102.5 | 28.8 | 8.73 | 4.43 | 0.085 | 0.154 |
| | Simulated/observed | 0.546 | 0.702 | 0.597 | 0.578 | 1.11 | 0.822 |

$AUC_{inf}$, area under the plasma concentration–time curve extrapolated to infinity; CI, confidence interval; $C_{max}$, maximum plasma concentration; CV%, coefficient of variation; $IndC_{50}$, calibrated inducer concentration producing half maximal induction; $Ind_{max}$, calibrated maximal fold induction over vehicle control.

The figure below shows the simulated (lines) and observed (circles; n = 19; DNDi-OXA-07-HAT) plasma concentration–time profiles of midazolam following a single oral dose of midazolam (5 mg) in the absence of acoziborole (upper panels) and on the 10th day post acoziborole single 960 mg dose (lower panels). Lines are the arithmetic mean data of the simulated population (90% CI) (n = 190).

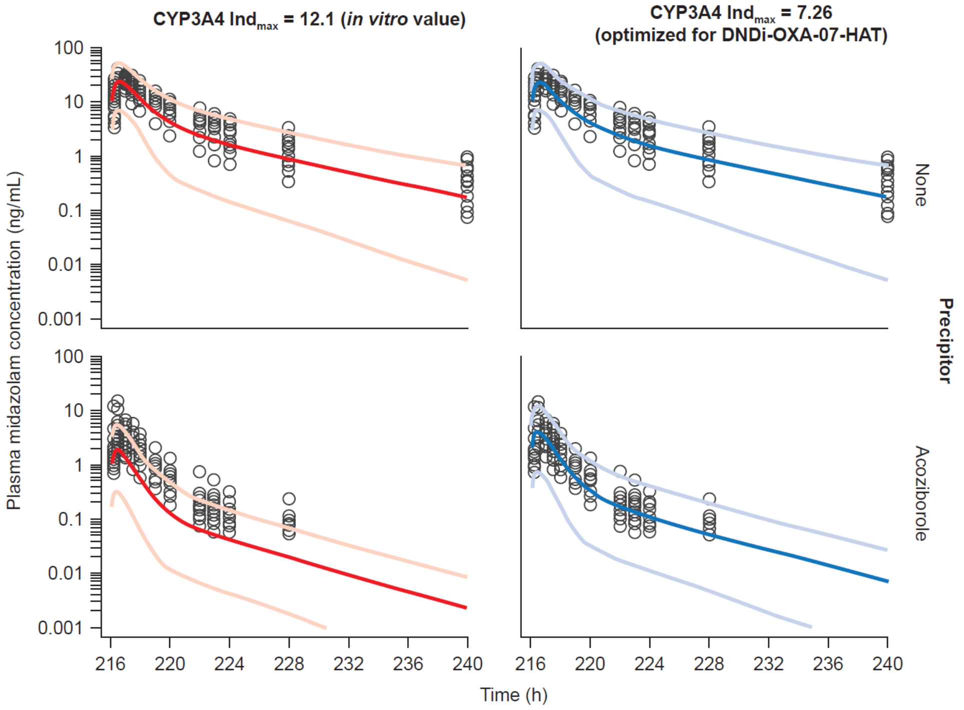

The figure below shows simulated enzyme-abundance profiles of CYP3A4 in the colon and small intestine (blue) and the liver (green) as a function of time following a single 960 mg dose of acoziborole (time-dependent CYP3A4 enzyme induction). The lines represent the arithmetic mean data for the simulated population (n = 190).

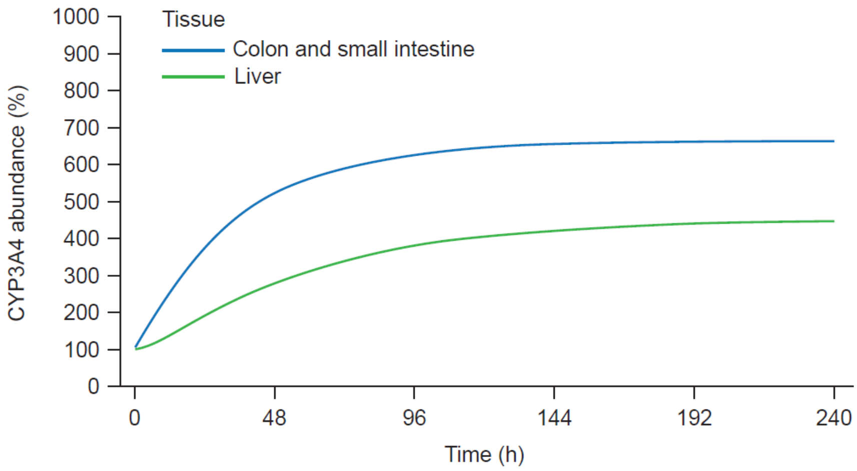

***CYP2S6 inhibition: Simulation of single oral dose of 15 mg dextromethorphan co-administered with a single oral dose of 960 mg acoziborole in healthy participants.***

Using the model above with refined induction parameters, the *in vitro* CYP2D6 inactivation parameters ($k_{inact}$, 4.46 $h^{-1}$; and $K_{app}$, 19.2 µM) underpredicted the observed geometric mean values for $AUC_t$, $AUC_{24}$ and $C_{max}$ for dextromethorphan in clinical study DNDi-OXA-07-HAT (table below). Using a sensitivity analysis, $k_{inact}$ was refined to 89.2 $h^{-1}$ with $K_{app}$ fixed at 19.2 µM.

| Model | Population | Without acoziborole | | | With acoziborole | | | Geometric mean ratio | | |
|---|---|---|---|---|---|---|---|---|---|---|
| | | $AUC_t$ (ng/mL·h) | $AUC_{24}$ (ng/mL·h) | $C_{max}$ (ng/mL) | $AUC_t$ (ng/mL·h) | $AUC_{24}$ (ng/mL·h) | $C_{max}$ (ng/mL) | $AUC_t$ | $AUC_{24}$ | $C_{max}$ |
| $k_{inact}$ 4.46 $h^{-1}$ | Simulated | 10.8 | 9.43 | 1.28 | 194 | 140 | 13.2 | 18.0 | 14.8 | 10.3 |
| | CV% | 75.7 | 75.7 | 73.2 | 55.3 | 46.5 | 33.6 | 47.9 | 47.8 | 50.0 |
| | 90% CI | 9.94, 11.7 | 8.7, 10.2 | 1.18, 1.38 | 182, 206 | 132, 147 | 12.6, 13.7 | 17.0, 19.0 | 14.0, 15.6 | 9.73, 10.9 |
| | Observed | 10.5 | 10.1 | 0.948 | 370 | 208 | 12.3 | 35.3 | 20.5 | 13.0 |
| | Simulated/observed | 1.03 | 0.932 | 1.35 | 0.523 | 0.673 | 1.07 | 0.510 | 0.720 | 0.792 |
| $k_{inact}$ 89.2 $h^{-1}$ | Simulated | 10.8 | 9.43 | 1.28 | 344 | 218 | 17.9 | 31.9 | 23.1 | 14.0 |
| | CV% | 75.7 | 75.7 | 73.2 | 47.0 | 35.3 | 25.5 | 73.7 | 69.3 | 67.4 |
| | 90% CI | 9.94, 11.7 | 8.70, 10.2 | 1.18, 1.38 | 326, 363 | 209, 227 | 17.3, 18.4 | 29.5, 34.6 | 21.4, 24.9 | 13.0, 15.1 |
| | Observed | 10.5 | 10.1 | 0.948 | 370 | 208 | 12.3 | 35.3 | 20.5 | 13.0 |
| | Simulated/observed | 1.03 | 0.934 | 1.35 | 0.930 | 1.05 | 1.45 | 0.905 | 1.12 | 1.08 |

$AUC_t$, area under the plasma concentration–time curve until the last quantifiable concentration; $AUC_{24}$, AUC until 24 h post-dose; $C_{max}$, maximum plasma concentration; CI, confidence interval; CV%, coefficient of variation; $K_{app}$, apparent inactivation constant (concentration supporting half-maximal rate of time-dependent inhibition); $k_{inact}$, maximal rate constant for enzyme inactivation.

The figure below shows the simulated (lines) and observed (circles; n = 19; DNDi-OXA-07-HAT) plasma concentration–time profiles of dextromethorphan following a single oral dose of dextromethorphan (15 mg) in the absence of acoziborole (upper panels) and on the 3rd day post acoziborole single 960 mg dose (lower panels). Lines are the arithmetic mean data of the simulated population (90% CI) (n = 190).

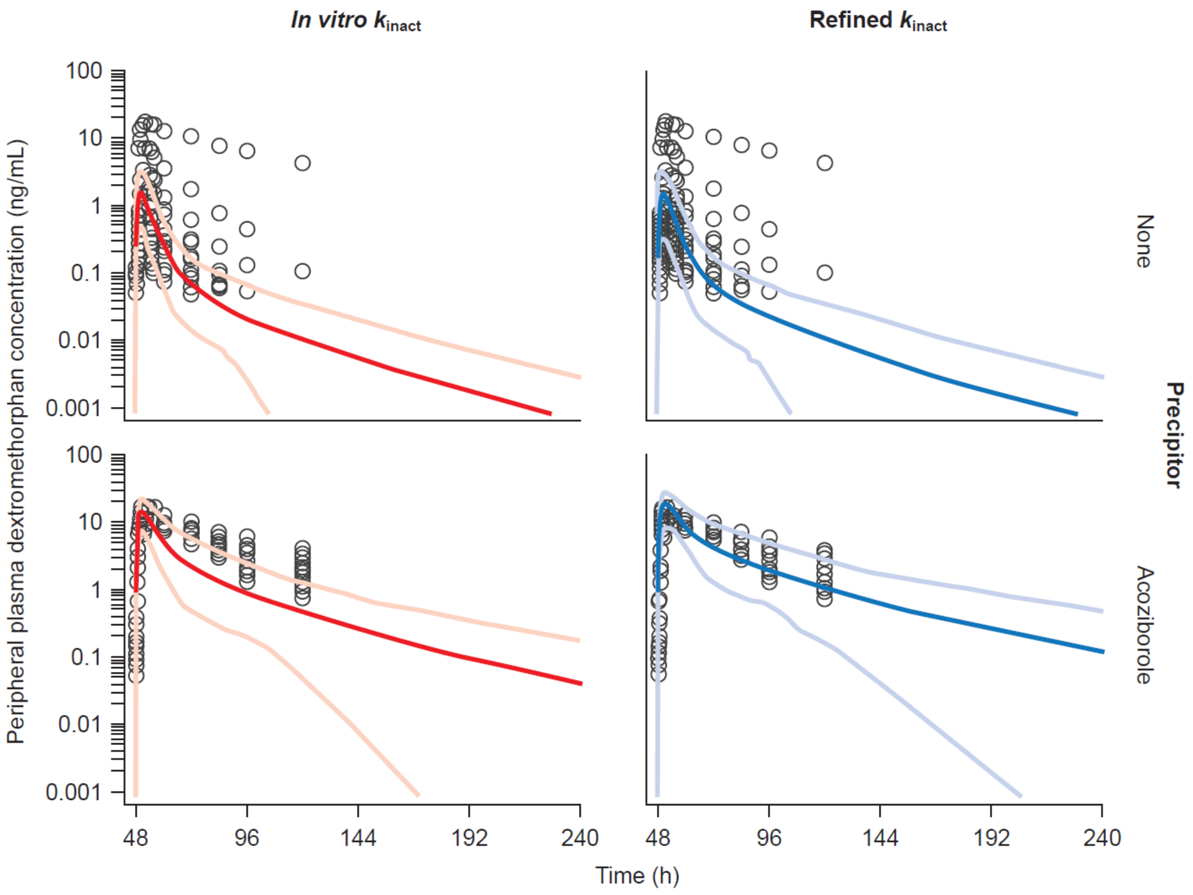

The figure below shows the simulated enzyme-abundance profiles of CYP2D6 following a single 960 mg dose of acoziborole (green: CYP2D6 in liver; blue: CYP2D6 in colon and small intestine). The lines represent the arithmetic mean data for the simulated population (n = 190). The simulated population was enriched with CYP2D6 extensive metabolizers.

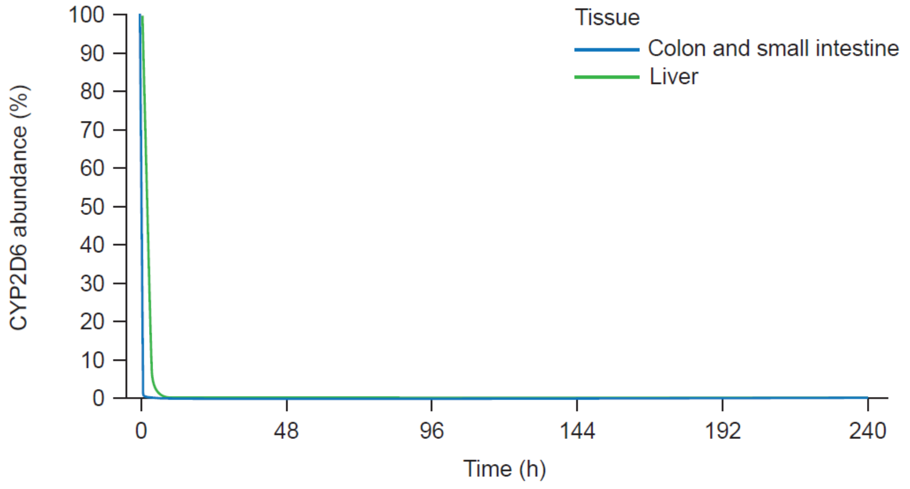

**Table S2: Definitions of strong, moderate and weak interactions as per the ICH M12 guideline on drug interaction studies**

| | | |
|---|---|---|
| Induction | Strong | decreases the AUC of a sensitive index CYP substrate by ≥ 80% |
| | Moderate | decreases the AUC of a sensitive index CYP substrate by ≥ 50% to < 80% |
| | Weak | decreases the AUC of a sensitive index CYP substrate by ≥ 20% to < 50% |
| Inhibition | Strong | increases the AUC of a sensitive index CYP substrate ≥ 5-fold |
| | Moderate | increases the AUC of a sensitive index CYP substrate ≥ 2- to < 5-fold |
| | Weak | increases the AUC of a sensitive index CYP substrate ≥ 1.25- to < 2-fold |

**Table S3: Dosing regimens of CYP3A4 substrate co-medications (objects of drug-drug interactions) and simulation designs used for acoziborole as drug-drug interaction precipitant.**

| Co-medication | Class | Dose regimen | Fraction metabolized / pathways | Object dosing day | Trial duration (days) |
|---|---|---|---|---|---|
| Lumefantrine | Anti-malarial | Standard regimen: 480 mg BID (6 doses) | CYP3A4 – 40%<br>Additional HLM – 60% | 10–12 | 28 |
| | | Extended regimen: 480 mg BID (11 doses) | CYP3A4 – 40%<br>Additional HLM – 60% | 10–15 | 28 |
| | | Standard regimen: 480 mg BID (6 doses) | CYP3A4 – 40%<br>Additional HLM – 60% | 36–38 | 54 |
| | | Extended regimen: 480 mg BID (11 doses) | CYP3A4 – 40%<br>Additional HLM – 60% | 36–41 | 54 |
| Artemether | | Standard regimen: 80 mg BID (6 doses) | CYP3A4 – 79%<br>CYP2B6 – 21% | 10–12 | 28 |
| | | Standard regimen: 80 mg BID (6 doses) | CYP3A4 – 79%<br>CYP2B6 – 21% | 36–38 | 54 |
| | | Extended regimen: 80 mg BID (11 doses) | CYP3A4 – 79%<br>CYP2B6 – 21% | 36–41 | 54 |
| Lopinavir/ritonavir | Anti-HIV | 800/200 mg QD (10 doses) | CYP3A4 – 99.8%<br>Renal – 0.2% | 1–10 | 10 |
| Dolutegravir | | 50 mg QD (10 doses) | CYP3A4 – 25%<br>UGT1A1 – 47%<br>Additional HLM – 28% | 1–10 | 10 |
| Ethinyl estradiol | Oral contraceptive | 0.035 mg QD (22 doses) | CYP3A4 – 23.35%<br>UGT1A1 – 5.47%<br>CYP1A2 – 8.42%<br>CYP2C8 – 0.85%<br>CYP2C9 – 11.34%<br>Additional HLM – 39.21%<br>Renal – 11.36% | 10–31 | 31 |

[a] Acoziborole was dosed on Day 1 (t = 0 h).
All co-medication simulations were dosed orally.
BID, twice a day; CYP, cytochrome P450; HIV, human immunodeficiency virus; HLM, human liver microsomes; QD, once a day; UGT, UDP-glucuronosyltransferase enzyme.

**Table S4: Dosing regimens of sensitive probe substrates (objects of drug-drug interactions) used for acoziborole as drug-drug interaction precipitant.**

| Probe substrate | Dose regimen | Fraction metabolized /pathways | Object dosing[a] | Trial duration, h |
|---|---|---|---|---|
| Caffeine | 150 mg | CYP1A2 – 98.3%<br>CYP2E1 – 0.71%<br>CYP3A4 – 0.30%<br>Renal elimination – 0.69% | 165 h<br>(Day 8 06:00) | 309 |
| Bupropion | 130.2 mg | CYP2B6 – 47.5%<br>CYP2C19 – 3.5%<br>CYP3A4 – 3.0%<br>Other hepatic metabolism – 43.9%<br>Renal elimination – 2.2% | 173 h<br>(Day 8 14:00) | 317 |
| Repaglinide | 0.25 mg | CYP2C8 – 62.4%<br>CYP3A4 – 37.4%<br>Renal elimination – 0.2% | 192 h<br>(Day 9 09:00) | 198 |
| Rosiglitazone | 4 mg | CYP2C8 – 56.2%<br>CYP2C9 – 27.4%<br>Other hepatic metabolism – 16.5% | 187 h<br>(Day 9 04:00) | 259 |
| S-warfarin | 10 mg | CYP2C9 – 97.5%<br>Renal elimination – 2.5% | 648 h<br>(Day 28 09:00) | 912 |
| Omeprazole | 20 mg | CYP2C19 – 86.14%<br>CYP3A4 – 13.86% | 312 h<br>(Day 14 09:00) | 336 |
| Rosuvastatin | 20 mg | BCRP – major efflux<br>OATP1B1/3 and OATP2B1 – major hepatic uptake<br>NTCP – minor<br>MRP4 – minor | 67 h<br>(Day 4 04:00) | 211 |

[a] Time post acoziborole dosing (hours).
All probe substrate simulations were dosed orally.
BCRP, breast cancer resistance protein; CYP, cytochrome P450; OATP, organic anion transporting polypeptide; MRP, multi-drug resistance-associated protein; NTCP, $Na^+$/taurocholate co-transporting polypeptide.

Simulations were performed with a single oral dose of 960 mg acoziborole administered at 9 a.m. on day 1 with the timing of the object drug single-dose administration adjusted depending on the specific pathways involved in its disposition. For objects of solely competitive inhibition by acoziborole, this meant administering the object and precipitant drugs such that they would have approximately overlapping $t_{max}$ values, and these object drugs were administered on day 4 at 4 a.m. This was the case for the BCRP substrate rosuvastatin. For interactions with time-dependent changes in pharmacokinetics, the time of object drug dosing varied, depending on the turnover of the enzymes involved to generate the maximum interaction (see above).

**Figure S3: Model verification – simulated concentration–time profiles for acoziborole compared with observed data.**

A. Simulated plasma concentration–time profile in healthy African participants and observed clinical data after a single 800 mg acoziborole dose (DNDi-OXA-001).
B. Simulated plasma concentration–time profile in healthy African participants and observed clinical data after a single 1000 mg acoziborole dose (DNDi-OXA-001).
C. Simulated blood concentration–time profile in adolescent African participants with g-HAT and observed clinical data after a single 960 mg acoziborole dose (DNDi-OXA-02-HAT).

The black line is the mean data for the simulated population; grey lines represent the 5th and 95th percentiles for the simulated population; open circles are observed data. Log–linear plots are shown on the left and linear plots on the right.

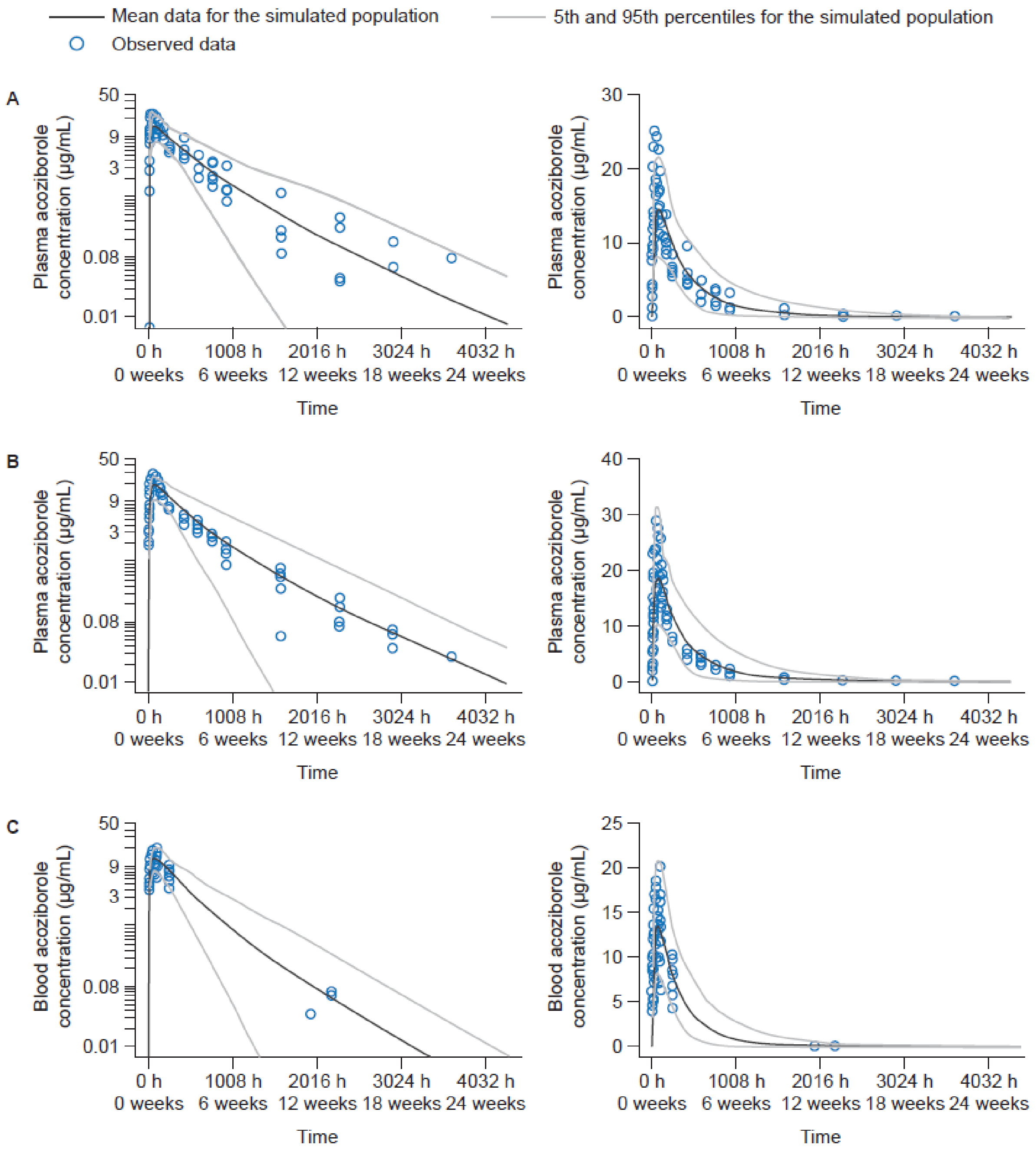

**Table S5: Model verification – simulated pharmacokinetic parameters for acoziborole independently verified against clinical data sets.**

***A) Healthy African participants: single oral dose of 800 or 1000 mg acoziborole (DNDi-OXA-001).***

| Dose | Statistic | $AUC_{inf}$ (µg/mL·h) | CL/F (L/h) | $C_{max}$ (µg/mL) | $t_{max}$ (h) |
|---|---|---|---|---|---|
| 800 mg | Simulated | 6376 | 0.125 | 14.0 | 74.7 |
| | CV% | 42.1 | 42.1 | 34.0 | – |
| | 90% CI | 5843, 6956 | 0.115, 0.137 | 13.1, 15.1 | 46.0, 118 |
| | Observed | 5899 | 0.136 | 17.0 | 48.0 |
| | Simulated/observed | 1.08 | 0.923 | 0.826 | 1.56 |
| 1000 mg | Simulated | 7735 | 0.129 | 18.0 | 74.4 |
| | CV% | 41.0 | 41.0 | 35.6 | – |
| | 90% CI | 7105, 8421 | 0.119, 0.141 | 16.7, 19.4 | 40.8, 121 |
| | Observed | 6886 | 0.145 | 22.5 | 72.0 |
| | Simulated/observed | 1.12 | 0.890 | 0.799 | 1.03 |

Values are geometric mean, except $t_{max}$ which is the median. $AUC_{inf}$, area under the plasma concentration–time curve extrapolated to infinity; CI, confidence interval ($t_{max}$ row lists range); $C_{max}$, maximum plasma concentration; CV%, coefficient of variation.

***B) Adolescent African participants with g-HAT (DNDi-OXA-02-HAT).***

| Dose | Statistic | $AUC_{inf}$ (µg/mL·h) | CL/F (L/h) | $C_{max}$ (µg/mL) | $t_{max}$ (h) |
|---|---|---|---|---|---|
| 960 mg | Simulated | 5903 | 0.163 | 15.7 | 68.9 |
| | CV% | 42.8 | 42.8 | 29.4 | – |
| | 90% CI | 5226, 6193 | 0.155, 0.171 | 15.2, 16.3 | 41.6, 123 |
| | Observed | 5611 | 0.17 | 13.6 | 51.4[a] |
| | Simulated/observed | 1.05 | 0.951 | 1.16 | 1.34[a] |

Values are geometric mean, except $t_{max}$ which is the median. $AUC_{inf}$, area under the blood concentration–time curve extrapolated to infinity; CI, confidence interval ($t_{max}$ row lists range); $C_{max}$, maximum blood concentration; CV%, coefficient of variation.

[a]Available observed data was geometric mean for $t_{max}$ , so simulated/observed ratio is the median/geometric mean.

**Table S6: Dose staggering simulations of geometric mean (90% CI) plasma exposures for midazolam after a single oral dose of 5 mg midazolam with or without a single oral dose of 960 mg acoziborole in African adult participants with g-HAT.**

| Midazolam dosing[a] | Without acoziborole | | With acoziborole | | Geometric mean ratio | |
|---|---|---|---|---|---|---|
| | $AUC_{inf}$ (ng/mL·h) | $C_{max}$ (ng/mL) | $AUC_{inf}$ (ng/mL·h) | $C_{max}$ (ng/mL) | $AUC_{inf}$ ratio | $C_{max}$ ratio |
| Day 1 (0 weeks) | 25.7 (23.2, 28.6) | 14.5 (13.0, 16.1) | 24.9 (22.4, 27.7) | 14.3 (12.9, 16.0) | 0.968 (0.965, 0.971) | 0.989 (0.988, 0.990) |
| Day 2 (0 weeks) | 25.7 (23.2, 28.6) | 14.5 (13.0, 16.1) | 8.32 (7.39, 9.36) | 5.30 (4.67, 6.03) | 0.323 (0.305, 0.342) | 0.366 (0.348, 0.386) |
| Day 8 (1 week) | 25.7 (23.2, 28.6) | 14.5 (13.0, 16.1) | 2.97 (2.61, 3.39) | 2.08 (1.80, 2.40) | 0.115 (0.106, 0.126) | 0.143 (0.132, 0.156) |
| Day 15 (2 weeks) | 25.7 (23.2, 28.6) | 14.5 (13.0, 16.1) | 3.01 (2.64, 3.44) | 2.10 (1.82, 2.43) | 0.117 (0.107, 0.128) | 0.145 (0.133, 0.158) |
| Day 22 (3 weeks) | 25.7 (23.2, 28.6) | 14.5 (13.0, 16.1) | 3.48 (3.04, 3.98) | 2.39 (2.06, 2.76) | 0.135 (0.123, 0.148) | 0.165 (0.151, 0.180) |
| Day 29 (4 weeks) | 25.7 (23.2, 28.6) | 14.5 (13.0, 16.1) | 4.25 (3.69, 4.89) | 2.84 (2.45, 3.30) | 0.165 (0.149, 0.182) | 0.196 (0.179, 0.215) |
| Day 33 (4 weeks) | 25.7 (23.2, 28.6) | 14.5 (13.0, 16.1) | 4.81 (4.16, 5.56) | 3.17 (2.73, 3.69) | 0.187 (0.168, 0.207) | 0.219 (0.199, 0.241) |
| Day 36 (5 weeks) | 25.7 (23.2, 28.6) | 14.5 (13.0, 16.1) | 5.29 (4.56, 6.12) | 3.44 (2.96, 4.01) | 0.205 (0.185, 0.229) | 0.238 (0.216, 0.262) |
| Day 43 (6 weeks) | 25.7 (23.2, 28.6) | 14.5 (13.0, 16.1) | 6.56 (5.63, 7.64) | 4.17 (3.56, 4.87) | 0.255 (0.228, 0.285) | 0.288 (0.260, 0.318) |
| Day 50 (7 weeks) | 25.7 (23.2, 28.6) | 14.5 (13.0, 16.1) | 8.03 (6.87, 9.38) | 4.98 (4.26, 5.83) | 0.312 (0.279, 0.350) | 0.344 (0.310, 0.381) |
| Day 57 (8 weeks) | 25.7 (23.2, 28.6) | 14.5 (13.0, 16.1) | 9.64 (8.25, 11.3) | 5.86 (5.01, 6.86) | 0.375 (0.335, 0.419) | 0.405 (0.366, 0.449) |
| Day 64 (9 weeks) | 25.7 (23.2, 28.6) | 14.5 (13.0, 16.1) | 11.3 (9.69, 13.2) | 6.77 (5.80, 7.91) | 0.440 (0.395, 0.490) | 0.468 (0.423, 0.517) |
| Day 71 (10 weeks) | 25.7 (23.2, 28.6) | 14.5 (13.0, 16.1) | 13.0 (11.2, 15.1) | 7.67 (6.59, 8.92) | 0.505 (0.455, 0.560) | 0.530 (0.482, 0.582) |
| Day 85 (12 weeks) | 25.7 (23.2, 28.6) | 14.5 (13.0, 16.1) | 16.1 (13.9, 18.5) | 9.31 (8.08, 10.7) | 0.624 (0.571, 0.683) | 0.643 (0.592, 0.699) |
| Day 106 (15 weeks) | 25.7 (23.2, 28.6) | 14.5 (13.0, 16.1) | 19.6 (17.3, 22.3) | 11.2 (9.87, 12.7) | 0.764 (0.714, 0.818) | 0.775 (0.727, 0.826) |
| Day 127 (18 weeks) | 25.7 (23.2, 28.6) | 14.5 (13.0, 16.1) | 22.0 (19.6, 24.8) | 12.5 (11.1, 14.1) | 0.856 (0.815, 0.900) | 0.862 (0.823, 0.904) |

[a] Days post acoziborole dosing.

$AUC_{inf}$, area under the plasma concentration–time curve extrapolated to infinity; $C_{max}$, maximum plasma concentration.

| Interaction | Strong | Moderate | Weak | Negligible |
|---|---|---|---|---|
| | ≤0.20 | >0.20, ≤0.50 | >0.50, ≤0.80 | >0.80 |

**Table S7: Geometric mean plasma exposures for simulated lumefantrine drug-drug interactions with acoziborole or rifampicin in African participants with g-HAT.**

***Geometric mean, CV% and 90% CI of exposures with or without a single 960 mg dose of acoziborole.***

| Lumefantrine dosing[a] | Statistic | Standard regimen[b] without acoziborole | | Standard regimen[b] with acoziborole | | Standard regimen[b] geometric mean ratio | | Extended regimen[b] with acoziborole | |
|---|---|---|---|---|---|---|---|---|---|
| | | AUC[c] (ng/mL·h) | $C_{max}$ (ng/mL) | AUC[c] (ng/mL·h) | $C_{max}$ (ng/mL) | AUC[c] | $C_{max}$ | AUC[c] (ng/mL·h) | $C_{max}$ (ng/mL) |
| Day 10 | Simulated | 592402 | 9950 | 116936 | 2567 | 0.197 | 0.258 | 216535 | 2655 |
| | CV% | 44.3 | 30.9 | 75.9 | 60.1 | 45.8 | 40.2 | 75.9 | 61.4 |
| | 90% CI | 552217, 635511 | 9463, 10461 | 104553 to 130786 | 2341, 2814 | 0.184, 0.212 | 0.242, 0.275 | 193586, 242206 | 2417, 2916 |
| Day 36 | Simulated | 592402 | 9950 | 198402 | 3932 | 0.335 | 0.395 | 374940 | 4224 |
| | CV% | 44.3 | 30.9 | 81.3 | 62.8 | 55.4 | 45.5 | 81.4 | 64.0 |
| | 90% CI | 552217, 635511 | 9463, 10461 | 176275, 223307 | 3573, 4328 | 0.307, 0.365 | 0.368, 0.425 | 333095, 422040 | 3833, 4656 |

***Geometric mean, CV% and 90% CI of exposures with or without multiple rifampicin doses (600 mg once daily).***

| Lumefantrine dosing[d] | Statistic | Standard regimen[b] without rifampicin | | Standard regimen[b] with rifampicin | | Standard regimen[b] geometric mean ratio | | Extended regimen[b] with rifampicin | |
|---|---|---|---|---|---|---|---|---|---|
| | | AUC[c] (ng/mL·h) | $C_{max}$ (ng/mL) | AUC[c] (ng/mL·h) | $C_{max}$ (ng/mL) | AUC[c] | $C_{max}$ | AUC[c] (ng/mL·h) | $C_{max}$ (ng/mL) |
| Day 15 | Simulated | 590380 | 9910 | 117322 | 3040 | 0.199 | 0.307 | 216269 | 3093 |
| | CV% | 43.7 | 31.2 | 84.5 | 60.3 | 63.0 | 47.7 | 83.9 | 62.0 |
| | 90% CI | 550820, 632780 | 9421, 10425 | 103861, 132528 | 2772, 3335 | 0.181, 0.219 | 0.285, 0.331 | 191581, 244138 | 2814, 3401 |

[a] Days post acoziborole dosing.

[b] Standard regimen: 480 mg BID, 6 doses; Extended regimen: 480 mg BID, 11 doses.

[c] For day 10 dosing $AUC_{216\text{-}672h}$, and for day 36 $AUC_{840\text{-}1296h}$ with or without acoziborole and $AUC_{336\text{-}792h}$ with or without rifampicin.

[d] Lumefantrine was dosed on day 15 following rifampicin dose commencement.

AUC, area under the plasma concentration–time curve over the specified interval; CI, confidence interval; $C_{max}$, maximum plasma concentration; CV%, coefficient of variation.

| Interaction | Strong | Moderate | Weak | Negligible |
|---|---|---|---|---|
| | ≤0.20 | >0.20, ≤0.50 | >0.50, ≤0.80 | >0.80 |

**Figure S4: Plasma concentration–time profiles for simulated lumefantrine drug-drug interactions with acoziborole or rifampicin in African participants with g-HAT.**

Lumefantrine plasma concentrations are shown.
Standard regimen: 480 mg BID, 6 doses; Extended regimen: 480 mg BID, 11 doses.

Note that the standard regimen with acoziborole and extended regimen with acoziborole are superimposed for the first 3 days of lumefantrine dosing.

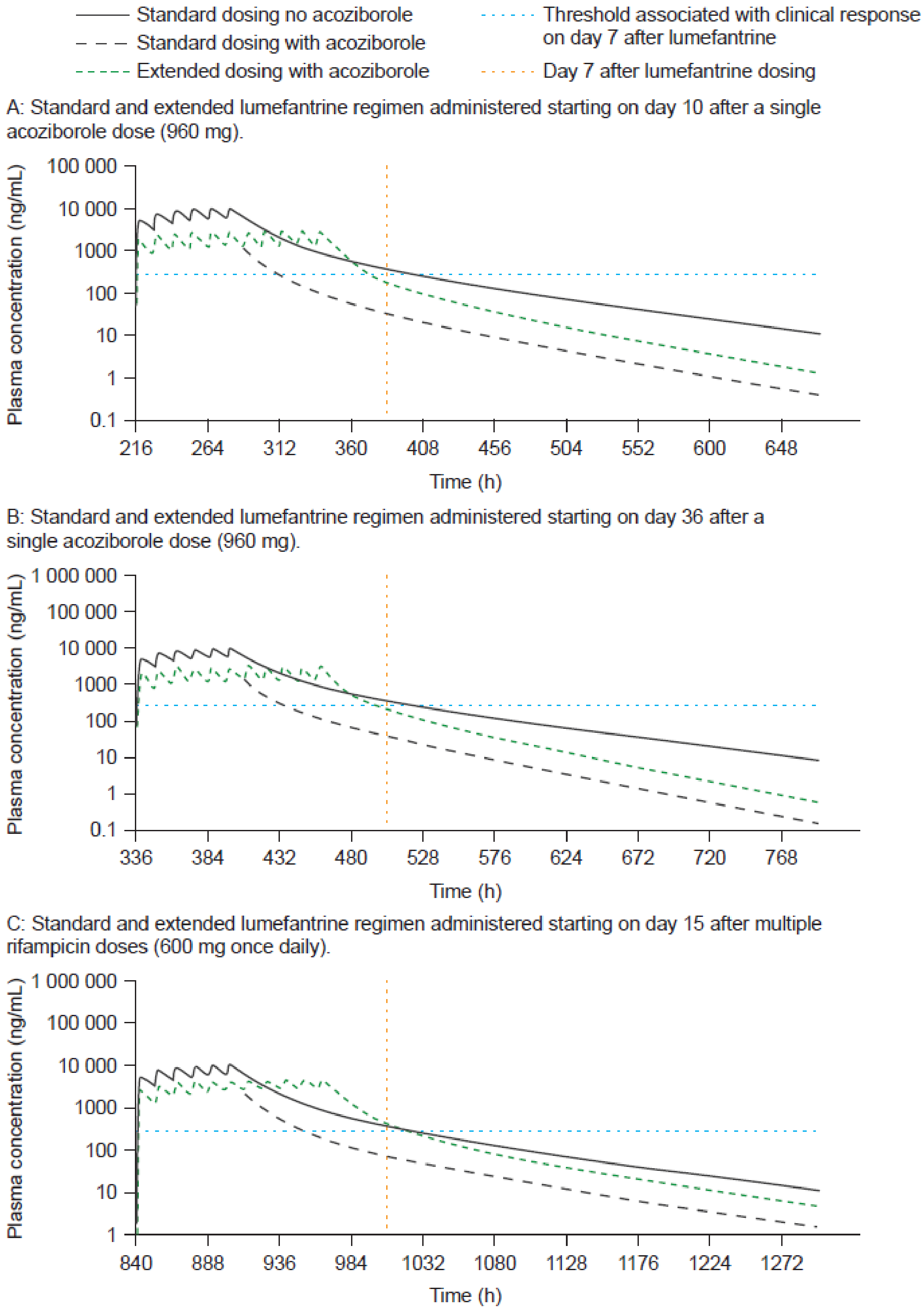

**Table S8: Geometric mean plasma exposures for simulated artemether drug-drug interactions with acoziborole or rifampicin in African participants with g-HAT.**

***Geometric mean, CV% and 90% CI of exposures with or without a single 960 mg dose of acoziborole.***

| Artemether dosing[a] | Statistic | Standard regimen[b] without acoziborole | | Standard regimen[b] with acoziborole | | Standard regimen[b] geometric mean ratio | | Extended regimen[b] with acoziborole | |
|---|---|---|---|---|---|---|---|---|---|
| | | AUC[c] (ng/mL·h) | $C_{max}$ (ng/mL) | AUC[c] (ng/mL·h) | $C_{max}$ (ng/mL) | AUC[c] | $C_{max}$ | AUC[c] (ng/mL·h) | $C_{max}$ (ng/mL) |
| Day 10 | Simulated | 1175 | 66.3 | 127 | 5.20 | 0.108 | 0.078 | NS | NS |
| | CV% | 52.5 | 53.6 | 84.7 | 92.7 | 65.6 | 55.2 | NS | NS |
| | 90% CI | 1082, 1275 | 61.0, 72.1 | 112, 143 | 4.56, 5.92 | 0.098, 0.119 | 0.072, 0.085 | NS | NS |
| Day 36 | Simulated | 1175 | 66.3 | 237 | 9.84 | 0.201 | 0.148 | 435 | 10.2 |
| | CV% | 52.5 | 53.6 | 98.1 | 108 | 76.9 | 74.5 | 94.6 | 105 |
| | 90% CI | 1082, 1275 | 61.0, 72.1 | 206, 271 | 8.51, 11.4 | 0.180, 0.225 | 0.133, 0.166 | 381, 497 | 8.82, 11.8 |

***Geometric mean, CV% and 90% CI of exposures with or without multiple rifampicin doses (600 mg once daily).***

| Artemether dosing[d] | Statistic | Standard regimen[b] without rifampicin | | Standard regimen[b] with rifampicin | | Standard regimen[b] geometric mean ratio | | Extended regimen[b] with rifampicin | |
|---|---|---|---|---|---|---|---|---|---|
| | | AUC[c] (ng/mL·h) | $C_{max}$ (ng/mL) | AUC[c] (ng/mL·h) | $C_{max}$ (ng/mL) | AUC[c] | $C_{max}$ | AUC[c] (ng/mL·h) | $C_{max}$ (ng/mL) |
| Day 15[b] | Simulated | 1150 | 65.9 | 188 | 13.2 | 0.163 | 0.201 | 347 | 13.2 |
| | CV% | 49.4 | 52.8 | 113 | 135 | 82.2 | 82.9 | 110 | 135 |
| | 90% CI | 1064, 1243 | 60.7, 71.6 | 161, 218 | 11.2, 15.7 | 0.145, 0.184 | 0.178, 0.227 | 299, 403 | 11.2, 15.7 |

[a] Days post acoziborole dosing.
[b] Standard regimen: 80 mg BID, 6 doses; Extended regimen: 80 mg BID, 11 doses.
[c] For day 10 dosing $AUC_{216\text{-}672h}$, and for day 36 $AUC_{840\text{-}1296h}$ with or without acoziborole and $AUC_{336\text{-}792h}$ with or without rifampicin.
[d] Artemether was dosed on day 15 following rifampicin dose commencement.
AUC, area under the plasma concentration–time curve over the specified interval; CI, confidence interval; $C_{max}$, maximum plasma concentration; CV%, coefficient of variation; NS, not simulated.

| Interaction | Strong | Moderate | Weak | Negligible |
|---|---|---|---|---|
| | ≤0.20 | >0.20, ≤0.50 | >0.50, ≤0.80 | >0.80 |

**Figure S5: Plasma concentration–time profiles for simulated artemether drug-drug interactions with acoziborole in African participants with g-HAT.**

Artemether plasma concentrations are shown.
Standard regimen: 80 mg BID, 6 doses; Extended regimen: 80 mg BID, 11 doses.

Standard and extended artemether regimen administered starting on day 36 after a single acoziborole dose (960 mg).

Note that the standard regimen with acoziborole and extended regimen with acoziborole are superimposed for the first 3 days of artemether dosing.

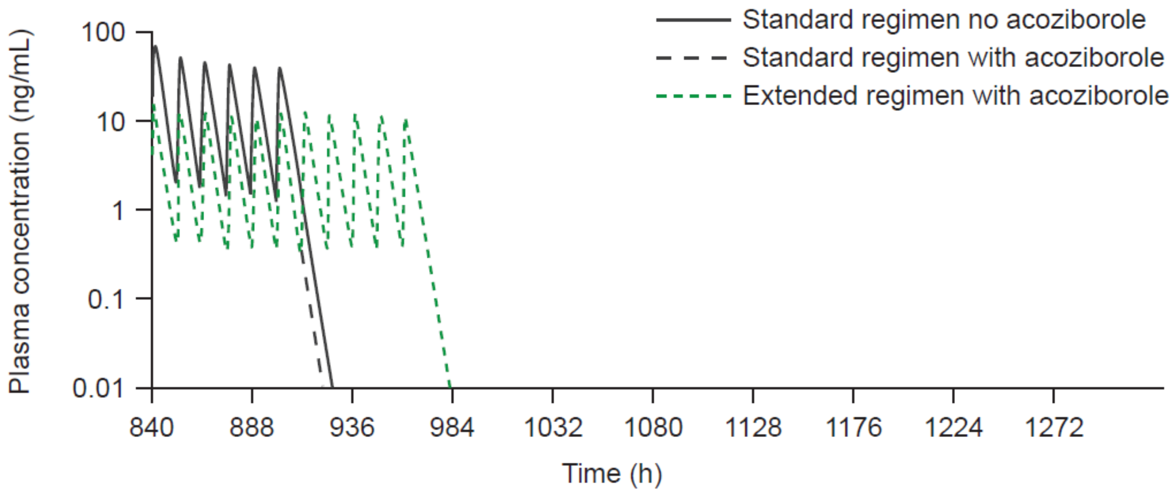

**Table S9: Geometric mean plasma exposures for simulated co-medication drug-drug interactions with acoziborole in African participants with g-HAT.** ***Geometric mean (90% CI) exposures with or without a single 960 mg dose of acoziborole.***

| Co-medication | Statistic | Without acoziborole | | With acoziborole | | Geometric mean ratio | |
|---|---|---|---|---|---|---|---|
| | | AUC[a] (ng/mL·h) | $C_{max}$ (ng/mL) | AUC[a] (ng/mL·h) | $C_{max}$ (ng/mL) | AUC[a] | $C_{max}$ |
| Lopinavir 800 mg QD (+ritonavir 200 mg QD) Multiple oral doses | Simulated | 76076 | 8089 | 30908 | 4240 | 0.406 | 0.524 |
| | CV% | 97.6 | 57.4 | 174 | 121 | 82.3 | 72.9 |
| | 90% CI | 66414, 87143 | 7403, 8838 | 25411, 37594 | 3620, 4966 | 0.361, 0.458 | 0.470, 0.584 |
| Dolutegravir 50 mg QD Multiple oral doses *In vitro* UGT1A1 $Ind_{max}$ | Simulated | 33088 | 2857 | 9066 | 1792 | 0.274 | 0.627 |
| | CV% | 45.5 | 32.8 | 59.0 | 36.0 | 27.4 | 12.4 |
| | 90% CI | 30789, 35559 | 2710, 3013 | 8279, 9928 | 1691, 1899 | 0.262, 0.287 | 0.614, 0.640 |
| Dolutegravir 50 mg QD Multiple oral doses UGT1A1 $Ind_{max}$ calibrated to rifampicin | Simulated | 33088 | 2857 | 15838 | 2164 | 0.479 | 0.757 |
| | CV% | 45.5 | 32.8 | 50.6 | 32.6 | 23.8 | 10.5 |
| | 90% CI | 30789, 35559 | 2710, 3013 | 14631, 17145 | 2053, 2281 | 0.460, 0.498 | 0.744, 0.771 |
| Ethinyl estradiol 0.035 mg QD Multiple oral doses *In vitro* UGT1A1 $Ind_{max}$ | Simulated | 16.2 | 0.0988 | 3.44 | 0.0326 | 0.212 | 0.330 |
| | CV% | 39.6 | 62.9 | 59.9 | 80.3 | 38.6 | 34.3 |
| | 90% CI | 15.3, 17.3 | 0.0898, 0.109 | 3.14, 3.78 | 0.0290, 0.0366 | 0.199, 0.226 | 0.312, 0.349 |
| Ethinyl estradiol 0.035 mg QD Multiple oral doses UGT1A1 $Ind_{max}$ calibrated to rifampicin | Simulated | 16.2 | 0.0988 | 5.16 | 0.0454 | 0.317 | 0.459 |
| | CV% | 39.6 | 62.9 | 56.1 | 76.1 | 34.8 | 28.5 |
| | 90% CI | 15.3, 17.3 | 0.0898, 0.109 | 4.73, 5.63 | 0.0405, 0.0507 | 0.300, 0.336 | 0.438, 0.481 |

[a] Lopinavir $AUC_{216\text{-}240h}$, dolutegravir $AUC_{216\text{-}240h}$, ethinyl estradiol $AUC_{216\text{-}744h}$.

AUC, area under the plasma concentration–time curve over the specified interval; CI, confidence interval; $C_{max}$, maximum plasma concentration; CV%, coefficient of variation.

| Interaction | Strong | Moderate | Weak | Negligible |
|---|---|---|---|---|
| | ≤0.20 | >0.20 to ≤0.50 | >0.50 to ≤0.80 | >0.80 |

**Figure S6: Plasma concentration–time profile for simulated ritonavir-boosted lopinavir drug-drug interactions with acoziborole in African participants with g-HAT.**

A: Plasma concentration–time profile (steady state) for lopinavir following multiple oral doses (800 mg QD) alone, boosted with ritonavir (lopinavir/ritonavir 800/200 mg QD) and lopinavir/ritonavir administered from day 10 after a single dose of acoziborole (960 mg).

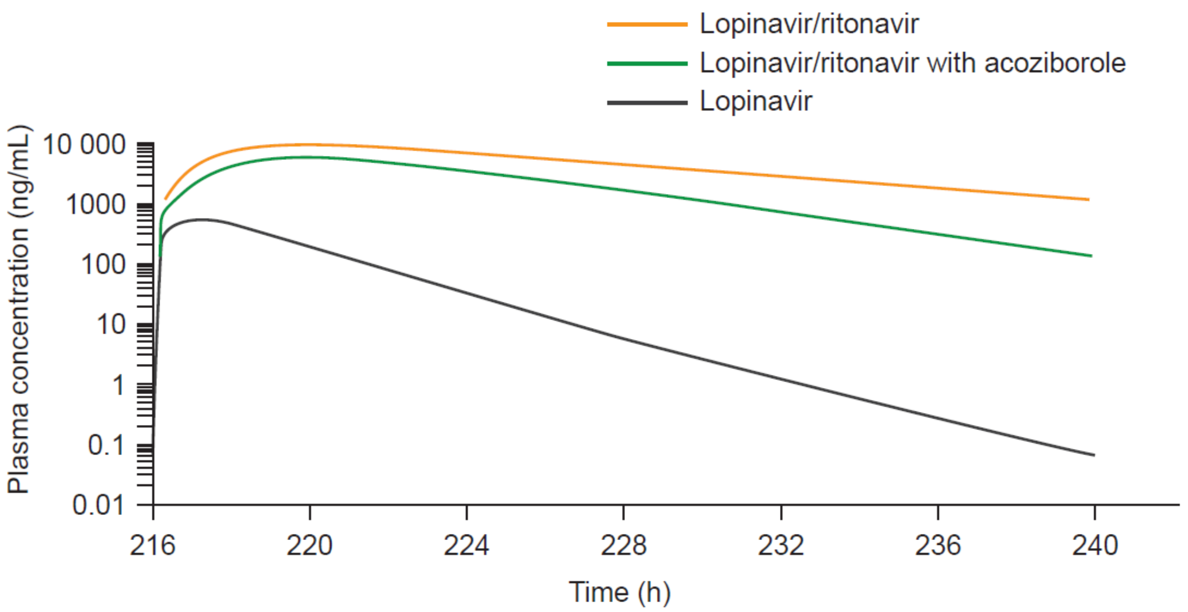

B: Flow chart illustrating the CYP3A4-mediated interactions affecting lopinavir when administered with ritonavir and acoziborole.

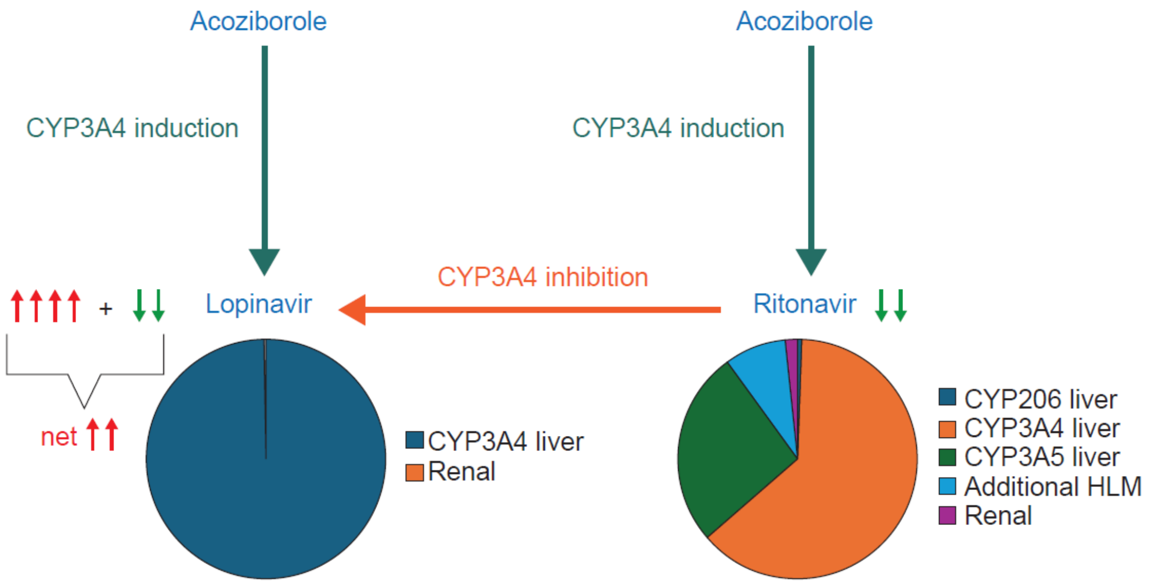

Arrows indicate effect on exposure (up = increase, down = decrease).

**Figure S7: Plasma concentration–time profile for simulated dolutegravir drug-drug interactions with acoziborole in African participants with g-HAT.**

Plasma concentration–time profile (steady state) for dolutegravir following multiple oral doses (50 mg QD) alone and administered from day 10 after a single dose of acoziborole (960 mg). UGT1A1 $Ind_{max}$ calibrated to rifampicin.

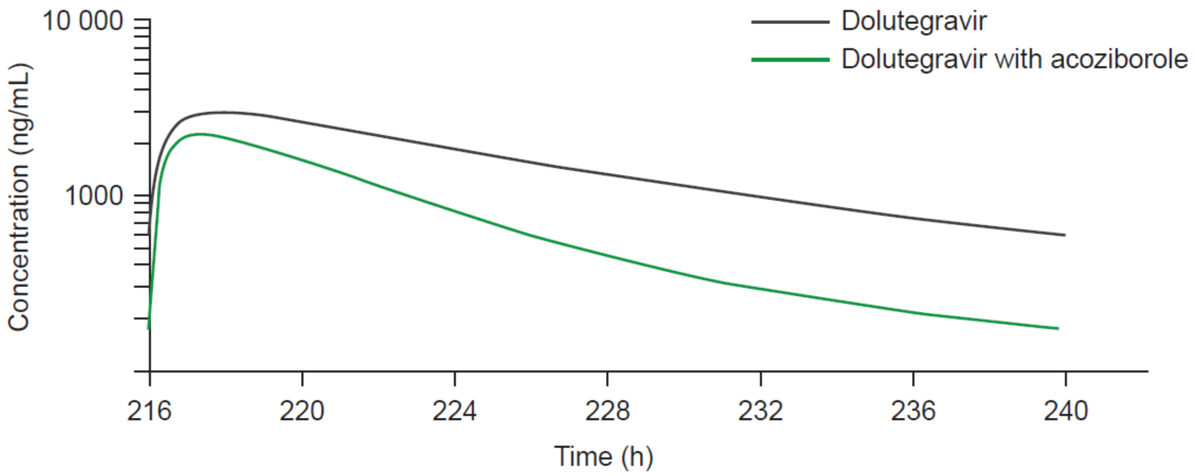

**Figure S8: Plasma concentration–time profiles for simulated ethinyl estradiol drug-drug interactions with acoziborole in African participants with g-HAT.**

Plasma concentration–time profile (steady state) for ethinyl estradiol following multiple oral doses (0.035 mg QD) alone and administered from day 10 after a single dose of acoziborole (960 mg). UGT1A1 $Ind_{max}$ calibrated to rifampicin.

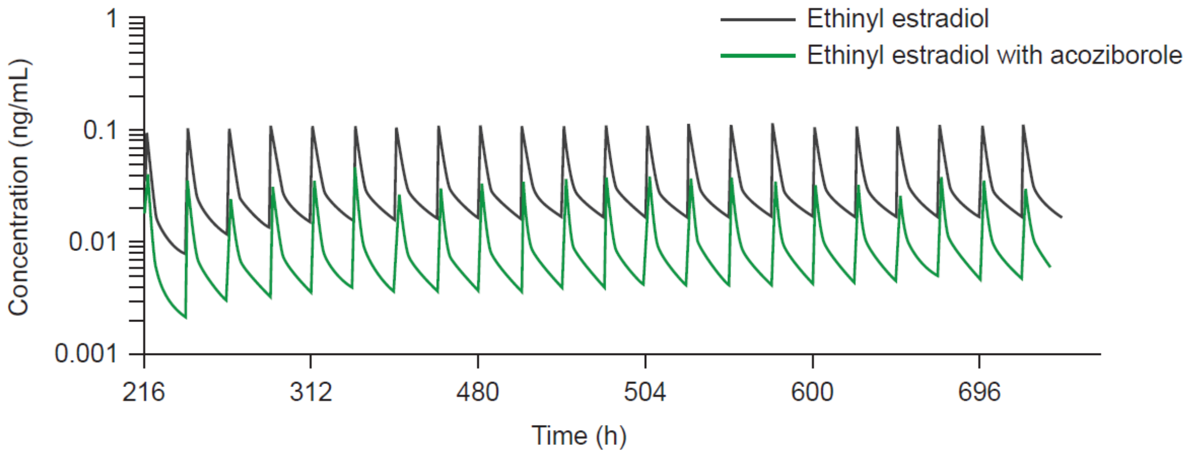

**Table S10: Dose staggering simulations of geometric mean (90% CI) plasma exposures for dextromethorphan after a single oral dose of 15 mg dextromethorphan with or without a single oral dose of 960 mg acoziborole in African adult participants with g-HAT.**

| Dextromethorphan dosing[a] | Without acoziborole | | With acoziborole | | Geometric mean ratio | |
|---|---|---|---|---|---|---|
| | $AUC_{inf}$ (ng/mL·h) | $C_{max}$ (ng/mL) | $AUC_{inf}$ (ng/mL·h) | $C_{max}$ (ng/mL) | $AUC_{inf}$ ratio | $C_{max}$ ratio |
| Day 1 (0 weeks) | 11.3 (9.83, 13.0) | 1.41 (1.25, 1.59) | 258 (237, 281) | 10.9 (10.2, 11.6) | 22.8 (20.6, 25.2) | 7.72 (7.04, 8.46) |
| Day 8 (1 week) | 11.3 (9.83, 13.0) | 1.41 (1.25, 1.59) | 267 (246, 290) | 17.5 (16.6, 18.4) | 23.6 (20.9, 26.7) | 12.4 (11.2, 13.8) |
| Day 29 (4 weeks) | 11.3 (9.83, 13.0) | 1.41 (1.25, 1.59) | 300 (276, 326) | 18.4 (17.5, 19.3) | 26.5 (23.9, 29.5) | 13.1 (11.8, 14.4) |
| Day 43 (6 weeks) | 11.3 (9.83, 13.0) | 1.41 (1.25, 1.59) | 244 (215, 278) | 16.0 (14.8, 17.4) | 21.6 (18.9, 24.7) | 11.4 (10.2, 12.7) |
| Day 64 (9 weeks) | 11.3 (9.83, 13.0) | 1.41 (1.25, 1.59) | 116 (95.4, 142) | 9.56 (8.26, 11.1) | 10.3 (8.55, 12.4) | 6.78 (5.87, 7.84) |
| Day 85 (12 weeks) | 11.3 (9.83, 13.0) | 1.41 (1.25, 1.59) | 55.0 (43.8, 69.0) | 5.35 (4.48, 6.37) | 4.87 (3.97, 5.97) | 3.79 (3.23, 4.46) |
| Day 92 (13 weeks) | 11.3 (9.83, 13.0) | 1.41 (1.25, 1.59) | 44.9 (35.7, 56.4) | 4.53 (3.78, 5.43) | 3.97 (3.24, 4.87) | 3.22 (2.74, 3.78) |
| Day 120 (17 weeks) | 11.3 (9.83, 13.0) | 1.41 (1.25, 1.59) | 25.1 (20.2, 31.2) | 2.81 (2.35, 3.36) | 2.22 (1.85, 2.66) | 1.99 (1.72, 2.32) |
| Day 127 (18 weeks) | 11.3 (9.83, 13.0) | 1.41 (1.25, 1.59) | 22.7 (18.4, 28.1) | 2.59 (2.17, 3.09) | 2.01 (1.69, 2.38) | 1.84 (1.59, 2.12) |
| Day 169 (24 weeks) | 11.3 (9.83, 13.0) | 1.41 (1.25, 1.59) | 15.6 (13.1, 18.6) | 1.89 (1.63, 2.20) | 1.38 (1.24, 1.54) | 1.34 (1.22, 1.48) |
| Day 183 (26 weeks) | 11.3 (9.83, 13.0) | 1.41 (1.25, 1.59) | 14.5 (12.3, 17.1) | 1.77 (1.54, 2.04) | 1.28 (1.17, 1.41) | 1.26 (1.16, 1.37) |

[a] Days post acoziborole dosing.

$AUC_{inf}$, area under the plasma concentration–time curve extrapolated to infinity; $C_{max}$, maximum plasma concentration.

| Interaction | Negligible | Weak | Moderate | Strong |
|---|---|---|---|---|
| | >0.80 to <1.25 | ≥1.25 to <2.0 | ≥2.0 to <5.0 | ≥5.0 |

**Table S11: Dose staggering simulations of geometric mean (90% CI) plasma exposures for metoprolol after a single oral dose of 100 mg metoprolol with or without a single oral dose of 960 mg acoziborole in African adult participants with g-HAT.**

| Metoprolol dosing[a] | Without acoziborole | | With acoziborole | | Geometric mean ratio | |
|---|---|---|---|---|---|---|
| | $AUC_{inf}$ (ng/mL·h) | $C_{max}$ (ng/mL) | $AUC_{inf}$ (ng/mL·h) | $C_{max}$ (ng/mL) | $AUC_{inf}$ ratio | $C_{max}$ ratio |
| Day 1 (0 weeks) | 967 (870, 1073) | 187 (174, 201) | 3358 (3103, 3634) | 251 (237, 266) | 3.47 (3.27, 3.69) | 1.34 (1.30, 1.38) |
| Day 8 (1 week) | 967 (870, 1073) | 187 (174, 201) | 1983 (1820, 2160) | 274 (260, 290) | 2.05 (1.86, 2.26) | 1.47 (1.39, 1.55) |
| Day 29 (4 weeks) | 967 (870, 1073) | 187 (174, 201) | 2706 (2495, 2935) | 312 (297, 328) | 2.80 (2.56, 3.06) | 1.67 (1.58, 1.76) |
| Day 43 (6 weeks) | 967 (870, 1073) | 187 (174, 201) | 3341 (3086, 3617) | 338 (323, 354) | 3.46 (3.17, 3.76) | 1.81 (1.71, 1.91) |
| Day 64 (9 weeks) | 967 (870, 1073) | 187 (174, 201) | 3111 (2815, 3438) | 326 (308, 345) | 3.22 (2.93, 3.54) | 1.74 (1.65, 1.84) |
| Day 85 (12 weeks) | 967 (870, 1073) | 187 (174, 201) | 2294 (2034, 2588) | 286 (267, 306) | 2.37 (2.14, 2.63) | 1.53 (1.45, 1.61) |
| Day 92 (13 weeks) | 967 (870, 1073) | 187 (174, 201) | 2072 (1827, 2350) | 273 (254, 293) | 2.14 (1.93, 2.38) | 1.46 (1.38, 1.54) |
| Day 120 (17 weeks) | 967 (870, 1073) | 187 (174, 201) | 1534 (1341, 1755) | 235 (217, 255) | 1.59 (1.43, 1.76) | 1.26 (1.19, 1.32) |
| Day 127 (18 weeks) | 967 (870, 1073) | 187 (174, 201) | 1457 (1274, 1666) | 229 (212, 248) | 1.51 (1.37, 1.66) | 1.23 (1.17, 1.29) |
| Day 169 (24 weeks) | 967 (870, 1073) | 187 (174, 201) | 1185 (1049, 1339) | 208 (192, 225) | 1.23 (1.14, 1.31) | 1.11 (1.07, 1.15) |
| Day 183 (26 weeks) | 967 (870, 1073) | 187 (174, 201) | 1132 (1007, 1274) | 204 (188, 220) | 1.17 (1.10, 1.24) | 1.09 (1.05, 1.12) |

[a] Days post acoziborole dosing.

$AUC_{inf}$, area under the plasma concentration–time curve extrapolated to infinity; $C_{max}$, maximum plasma concentration.

| Interaction | Negligible | Weak | Moderate | Strong |
|---|---|---|---|---|
| | >0.80 to <1.25 | ≥1.25 to <2.0 | ≥2.0 to <5.0 | ≥5.0 |

**Table S12: Dose staggering simulations of geometric mean ratios (90% CI) for hypothetical CYP2D6 substrates with fractional metabolism ($f_m$) of 25%, 50% or 75% after a single oral dose of 100 mg with or without a single oral dose of 960 mg acoziborole in African adult participants with g-HAT.**

| Substrate dosing[a] | $f_m$ 25% CYP2D6 geometric mean ratio | | $f_m$ 50% CYP2D6 geometric mean ratio | | $f_m$ 75% CYP2D6 geometric mean ratio | |
|---|---|---|---|---|---|---|
| | $AUC_{inf}$ ratio | $C_{max}$ ratio | $AUC_{inf}$ ratio | $C_{max}$ ratio | $AUC_{inf}$ ratio | $C_{max}$ ratio |
| Day 1 (0 weeks) | 1.20<br>1.18, 1.23 | 1.07<br>1.06, 1.08 | 1.59<br>1.53, 1.65 | 1.16<br>1.15, 1.18 | 2.80<br>2.64, 2.97 | 1.31<br>1.27, 1.34 |
| Day 8 (1 week) | 1.37<br>1.33, 1.42 | 1.23<br>1.21, 1.26 | 2.16<br>2.03, 2.29 | 1.59<br>1.53, 1.66 | 4.77<br>4.36, 5.22 | 2.22<br>2.09, 2.35 |
| Day 29 (4 weeks) | 1.37<br>1.33, 1.41 | 1.23<br>1.20, 1.25 | 2.13<br>2.00, 2.26 | 1.58<br>1.52, 1.64 | 4.57<br>4.19, 5.00 | 2.18<br>2.06, 2.31 |
| Day 43 (6 weeks) | 1.35<br>1.31, 1.39 | 1.22<br>1.19, 1.24 | 2.03<br>1.92, 2.16 | 1.54<br>1.48, 1.60 | 4.06<br>3.70, 4.45 | 2.08<br>1.96, 2.20 |
| Day 64 (9 weeks) | 1.29<br>1.25, 1.32 | 1.18<br>1.16, 1.20 | 1.79<br>1.69, 1.90 | 1.43<br>1.38, 1.49 | 3.03<br>2.74, 3.34 | 1.82<br>1.71, 1.93 |
| Day 85 (12 weeks) | 1.20<br>1.17, 1.24 | 1.13<br>1.11, 1.15 | 1.53<br>1.44, 1.62 | 1.30<br>1.26, 1.35 | 2.20<br>1.98, 2.44 | 1.55<br>1.46, 1.64 |
| Day 92 (13 weeks) | 1.18<br>1.15, 1.21 | 1.12<br>1.10, 1.14 | 1.46<br>1.37, 1.55 | 1.27<br>1.22, 1.32 | 2.00<br>1.81, 2.22 | 1.47<br>1.39, 1.56 |
| Day 120 (17 weeks) | 1.11<br>1.08, 1.14 | 1.07<br>1.05, 1.09 | 1.26<br>1.20, 1.33 | 1.16<br>1.12, 1.20 | 1.51<br>1.38, 1.65 | 1.27<br>1.20, 1.33 |
| Day 127 (18 weeks) | 1.10<br>1.07, 1.12 | 1.06<br>1.05, 1.08 | 1.23<br>1.17, 1.29 | 1.14<br>1.10, 1.18 | 1.43<br>1.32, 1.56 | 1.23<br>1.17, 1.29 |
| Day 169 (24 weeks) | 1.05<br>1.03, 1.06 | 1.03<br>1.02, 1.04 | 1.10<br>1.06, 1.14 | 1.06<br>1.04, 1.09 | 1.17<br>1.11, 1.24 | 1.10<br>1.06, 1.14 |
| Day 183 (26 weeks) | 1.03<br>1.02, 1.05 | 1.02<br>1.01, 1.03 | 1.07<br>1.04, 1.11 | 1.05<br>1.03, 1.07 | 1.13<br>1.07, 1.18 | 1.08<br>1.04, 1.11 |

[a] Days post acoziborole dosing.

$AUC_{inf}$, area under the plasma concentration–time curve extrapolated to infinity; $C_{max}$, maximum plasma concentration.

| Interaction | Negligible | Weak | Moderate | Strong |
|---|---|---|---|---|
| | >0.80 to <1.25 | ≥1.25 to <2.0 | ≥2.0 to <5.0 | ≥5.0 |

**Table S13: Simulated geometric mean (90% CI) exposures for sensitive probe substrates with or without a single 960 mg dose of acoziborole in African participants with g-HAT.**

| Pathway | DDI object | Without acoziborole | | With acoziborole | | Geometric mean ratio | |
|---|---|---|---|---|---|---|---|
| | | AUC (ng/mL·h) | $C_{max}$ (ng/mL) | AUC (ng/mL·h) | $C_{max}$ (ng/mL) | AUC ratio | $C_{max}$ ratio |
| CYP1A2 | caffeine | 27262 (24526, 30303) | 4163 (3988, 4345) | 19970 (18051, 22094) | 3870 (3700, 4048) | 0.733 (0.717, 0.749) | 0.930 (0.924, 0.935) |
| CYP2B6 | bupropion | 1149 (1071, 1233) | 79.1 (74.9, 83.7) | 657 (607, 711) | 51.9 (48.4, 55.5) | 0.572 (0.556, 0.588) | 0.655 (0.640, 0.671) |
| | OH-bupropion | 10412 (9559, 11342) | 251 (231, 272) | 18223 (17149, 19365) | 481 (456, 508) | 1.75 (1.67, 1.83) | 1.92 (1.83, 2.01) |
| CYP2C8 | rosiglitazone | 1096 (1005, 1195) | 266 (255, 277) | 497 (448, 551) | 194 (183, 206) | 0.454 (0.439, 0.469) | 0.731 (0.713, 0.748) |
| CYP2C8 | repaglinide | 4.23 (3.74, 4.78) | 3.44 (3.07, 3.86) | 1.46 (1.29, 1.65) | 1.38 (1.21, 1.57) | 0.345 (0.328, 0.363) | 0.400 (0.382, 0.419) |
| CYP2C9 | S-warfarin | 41402 (36205, 47345) | 1385 (1319, 1455) | 20306 (17506, 23553) | 1281 (1219, 1346) | 0.490 (0.467, 0.515) | 0.925 (0.916, 0.933) |
| CYP2C19 | omeprazole | 376 (336, 422) | 171 (157, 187) | 123 (110, 136) | 67.8 (61.5, 74.8) | 0.325 (0.310, 0.341) | 0.396 (0.380, 0.413) |
| BCRP | rosuvastatin | 115 (99.2, 132) | 10.2 (8.83, 11.8) | 143 (125, 164) | 13.5 (11.7, 15.5) | 1.25 (1.23, 1.27) | 1.32 (1.29, 1.34) |

AUC, area under the concentration-time curve – for repaglinide, rosiglitazone, and omeprazole AUC extrapolated to infinity ($AUC_{inf}$) was used and for caffeine, bupropion, OH-bupropion, rosuvastatin, and S-warfarin, AUC until the time of last quantifiable concentration was used ($AUC_t$). $C_{max}$, maximum plasma concentration.

| Interaction | Strong | Moderate | Weak | Negligible | Weak | Moderate | Strong |
|---|---|---|---|---|---|---|---|
| | ≤0.20 | >0.20 to ≤0.50 | >0.50 to ≤0.80 | >0.80 to <1.25 | ≥1.25 to <2.0 | ≥2.0 to <5.0 | ≥5.0 |

**Table S14: Sensitivity analysis for simulated geometric mean (90% CI) exposures for sensitive probe substrates with or without a single 960 mg dose of acoziborole in African participants with g-HAT.**

| Pathway | DDI object | Sensitivity analysis | Geometric mean ratio | |
|---|---|---|---|---|
| | | | AUC ratio | $C_{max}$ ratio |
| CYP1A2 | caffeine | *In vitro* values | 0.733 (0.717, 0.749) | 0.930 (0.924, 0.935) |
| | | CYP1A2 $Ind_{max}$ increased x10 | 0.166 (0.157, 0.176) | 0.520 (0.501, 0.540) |
| CYP2B6 | bupropion | *In vitro* values | 0.572 (0.556, 0.588) | 0.655 (0.640, 0.671) |
| | | CYP2B6 $Ind_{max}$ increased x10 | 0.152 (0.140, 0.165) | 0.209 (0.193, 0.227) |
| | OH-bupropion | *In vitro* values | 1.75 (1.67, 1.83) | 1.92 (1.83, 2.01) |
| | | CYP2B6 $Ind_{max}$ increased x10 | 2.75 (2.58, 2.93) | 3.32 (3.11, 3.54) |
| CYP2C8/ CYP3A4 | repaglinide | *In vitro* values | 0.345 (0.328, 0.363) | 0.400 (0.382, 0.419) |
| | | No CYP3A4 induction | 0.712 (0.697, 0.726) | 0.777 (0.763, 0.791) |
| | | No CYP2C8 induction | 0.384 (0.363, 0.406) | 0.434 (0.413, 0.457) |
| | | CYP2C8 $K_i$ decreased x10 | 0.366 (0.347, 0.385) | 0.419 (0.400, 0.439) |
| | | No CYP2C8 inhibition | 0.343 (0.326, 0.360) | 0.397 (0.379, 0.417) |
| | | CYP2C8 $Ind_{max}$ increased x10 | 0.246 (0.232, 0.260) | 0.300 (0.285, 0.316) |
| CYP2C8 | rosiglitazone | *In vitro* values | 0.454 (0.439, 0.469) | 0.731 (0.713, 0.748) |
| | | No CYP2C8 induction | 0.829 (0.812, 0.847) | 0.937 (0.928, 0.945) |
| | | CYP2C8 $K_i$ decreased x10 | 0.566 (0.550, 0.583) | 0.808 (0.793, 0.822) |
| | | No CYP2C9 induction | 0.521 (0.500, 0.542) | 0.775 (0.757, 0.794) |
| | | No CYP2C8 inhibition | 0.439 (0.424, 0.454) | 0.719 (0.701, 0.737) |
| | | CYP2C8 $Ind_{max}$ increased x10 | 0.0777 (0.0724, 0.0834) | 0.235 (0.218, 0.255) |
| CYP2C9 | S-warfarin | *In vitro* values | 0.490 (0.467, 0.515) | 0.925 (0.916, 0.933) |
| | | CYP2C9 $Ind_{max}$ 2.07 $IndC_{50}$ 0.534 μM | 0.734 (0.711, 0.759) | 0.968 (0.964, 0.972) |
| | | CYP2C9 $Ind_{max}$ increased x10 | 0.0458 (0.0431, 0.0486) | 0.482 (0.458, 0.508) |
| CYP2C19 | omeprazole | *In vitro* values | 0.326 (0.310, 0.341) | 0.396 (0.380, 0.413) |
| | | CYP2C19 $Ind_{max}$ 3.14 $IndC_{50}$ 3.36 μM | 0.479 (0.456, 0.504) | 0.561 (0.543, 0.580) |
| | | No CYP3A4 induction | 0.421 (0.398, 0.445) | 0.486 (0.462, 0.512) |
| | | CYP2C19 $Ind_{max}$ increased x10 | 0.0314 (0.0287, 0.0344) | 0.0414 (0.0375, 0.0457) |
| BCRP | rosuvastatin | *In vitro* values | 1.25 (1.23, 1.27) | 1.32 (1.29, 1.34) |
| | | BCRP $K_i$ decreased x15 | 2.10 (1.99, 2.22) | 2.75 (2.60, 2.91) |
| | | BCRP $K_i$ decreased x15 $fu_{gut}$ = fu | 1.22 (1.21, 1.24) | 1.32 (1.30, 1.34) |

| Pathway | DDI object | Sensitivity analysis | Geometric mean ratio | |
|---|---|---|---|---|
| | | | AUC ratio | $C_{max}$ ratio |
| | | $fu_{gut}$ = fu | 1.04 (1.04, 1.04) | 1.05 (1.04, 1.05) |

AUC, area under the concentration-time curve – for repaglinide, rosiglitazone, and omeprazole AUC extrapolated to infinity ($AUC_{inf}$) was used and for caffeine, bupropion, OH-bupropion, rosuvastatin, and S-warfarin, AUC until the time of last quantifiable concentration was used ($AUC_t$). $C_{max}$, maximum plasma concentration; CYP, cytochrome P450; BCRP, breast cancer resistance protein; fu, fraction unbound in plasma; $fu_{gut}$, fraction unbound in gut enterocytes; $IndC_{50}$, calibrated inducer concentration producing half maximal induction; $Ind_{max}$, calibrated maximal fold induction over vehicle control; $K_i$, reversible inhibition constant.

| Interaction | Strong | Moderate | Weak | Negligible | Weak | Moderate | Strong |
|---|---|---|---|---|---|---|---|
| | ≤0.20 | >0.20 to ≤0.50 | >0.50 to ≤0.80 | >0.80 to <1.25 | ≥1.25 to <2.0 | ≥2.0 to <5.0 | ≥5.0 |
